# Real-world effectiveness of molnupiravir and nirmatrelvir/ritonavir among COVID-19 community, highly vaccinated patients with high risk for severe disease: Evidence that both antivirals reduce the risk for disease progression and death

**DOI:** 10.1101/2023.02.09.23285737

**Authors:** D. Paraskevis, M. Gkova, K. Mellou, G. Gerolymatos, P. Psalida, K. Gkolfinopoulou, E.G. Kostaki, S. Loukides, A. Kotanidou, A. Skoutelis, E. Thiraios, G. Saroglou, D. Zografopoulos, E. Mossialos, T. Zaoutis, M. Gaga, S. Tsiodras, A. Antoniadou

## Abstract

Besides the significant benefits of vaccination against COVID-19, the risk of severe disease and death from COVID-19 among highly vulnerable populations remains of concern. Implementation of oral antiviral treatment has shown significant benefits for outpatients with high risk for severe disease, however, their effectiveness remains to be evaluated in real-life settings and in the presence of new Omicron subvariants. We aimed to investigate the effectiveness of molnupiravir and nirmatrelvir/ritonavir using a retrospective cohort design with outcomes hospital admission and death from COVID-19, in Greece. The effectiveness of each drug was estimated through a comparison of the antiviral’s recipients with an age-matched control group of non-recipients, adjusted for age, previous SARS-CoV-2 infection, vaccination status, and vaccination recency. Our analysis showed that molnupiravir significantly reduced the risk for hospitalization (OR = 0.40, p < 0.001) and death from COVID-19 (OR = 0.31, p < 0.001), with the effect being more intense among elderly patients (≥75 years old). The effectiveness was higher among those with full adherence. Nirmatrelvir/ritonavir was found also to significantly reduce the risk of hospital admission (OR = 0.31, p < 0.001) and death (OR = 0.28, p < 0.001) and, similarly to molnupiravir, effectiveness was stronger among elderly patients and those with the highest levels of adherence. Analysis of the relative effectiveness of nirmatrelvir/ritonavir versus molnupiravir suggested that nirmatrelvir/ritonavir was associated with a reduced risk for hospital admission (OR = 0.58, p < 0.001) compared to molnupiravir, adjusted for age, previous SARS-CoV-2 infection, vaccination status, and co-morbidities. Our real-world study provides evidence about the reduced risk of hospitalization and death in highly vaccinated patients with a high risk for severe disease in Greece. These findings highlight that although the hospitalization and mortality risk has been reduced mainly due to vaccination and the emergence of Omicron variants, antivirals provide significant additional benefits in highly vulnerable patients and therefore their use is documented and strongly indicated.

## Introduction

Omicron variants of SARS-CoV-2 have been related to reduced severity of illness and risk of death but increased incidence of breakthrough infections [1]. Primary and boosted vaccination against SARS-CoV-2 has also significantly reduced the risk for severe disease and death associated with COVID-19 [2-5], however, SARS-CoV-2 infection remains significant morbidity and mortality issue among the highly vulnerable populations including the elderly, immunocompromised people, and those with chronic diseases [6-8]. Development of antivirals against SARS-CoV-2 is crucial to reduce the risk of hospitalization, disease progression, and death among high-risk patients. Remdesivir was the first antiviral approved for emergency use to treat COVID-19 patients and it has been shown to prevent disease progression in hospitalized patients with moderate to severe disease and also in outpatients [9, 10]. Remdesivir is limited by the intravenous route necessary for its administration, while molnupiravir and nirmatrelvir/ritonavir are two newer agents administered orally that have shown in vitro and in vivo activity against SARS-CoV-2.

Molnupiravir use in outpatients has been shown to reduce the risk for hospitalization by 30% compared to placebo in a randomized, double-blind, phase 3 trial (MOVe-OUT Trial) [11]. Following these results, FDA issued an emergency use authorized (EUA) for molnupiravir for the treatment of patients with moderate-to-severe COVID-19, who have a high risk for disease progression, and for whom alternative FDA-authorized COVID-19 treatments are not accessible or appropriate [12].

Nirmatrelvir/ritonavir outpatient use has significantly reduced the risk for hospitalization and death (relative risk reduction, 88.9%) by day 28 after drug administration in symptomatic, unvaccinated, non-hospitalized patients at high risk for progression to severe COVID-19 [13]. Nirmatrelvir/ritonavir received conditional authorization in December 2021 in the United Kingdom, for the treatment of COVID-19 patients who are at high risk for severe disease and not requiring supplemental oxygen [14]. It also received emergency use authorization (EUA) in the USA for the treatment of mild-to-moderate COVID-19 patients at high risk for severe COVID-19 [15]. Both antivirals should be administered as soon as possible after SARS-CoV-2 diagnosis and within 5 days after symptoms onset. Nirmatrelvir/ritonavir subsequently received EU approval for use in January 2022, with the same indication as in the UK [16]. Molnupiravir and nirmatrelvir/ritonavir outpatient use in COVID-19 patients with a high risk for disease progression has shown evidence for a significant reduction in the risk for hospitalization and death in real-world studies, however, more real-world data is necessary [17-23].

We aimed to investigate the real-world effectiveness of molnupiravir, and nirmatrelvir/ritonavir administered in COVID-19 patients with a high risk for severe disease during a national government-audited program of administration in Greece between February and July 2022. The effectiveness of the antivirals was investigated through a comparison of the molnupiravir and nirmatrelvir/ritonavir recipients with age-matched non-recipients separately for each drug (drugs were not concomitantly but consecutively available in Greece).

## Patients and Methods

### Study population

The study population consisted of all SARS-CoV-2 infected non-hospitalized eligible patients who received: i) molnupiravir (800 mg twice daily for 5 days) between February 2, 2022 and March, 5, 2022 (N=6,474), and ii) nirmatrelvir/ritonavir (nirmatrelvir 300 mg and ritonavir 100 mg twice daily for 5 days, or nirmatrelvir 150 mg and ritonavir 100 mg twice daily, if estimated glomerular filtration rate was 30–59 mL/min per 1.73 m²) between March 26, 2022 and July 20, 2022 (N=23,191). Specifically, we selected all outpatient molnupiravir recipients aged 65 years or older (N= 4,240) corresponding to 65.5% of the total number of patients on molnupiravir and 13,861 nirmatrelvir/ritonavir recipients accounting for 59.8% of the total population treated with nirmatrelvir/ritonavir. We selected only the age group ≥ 65 years old, since co-morbidities were unknown among the non-recipients (controls). Given that co-morbidities increase with age, age-matched controls are very likely to have a similar pattern of co-morbidities as the antiviral recipients.

Antivirals were centrally available and dispensed to the public through hospital pharmacies and free courier service, while eligibility and prescribing requirements followed the national guidelines for SARS-CoV-2 antiviral treatment. Patients were eligible if they had a positive nucleic acid amplification test (NAAT), or rapid antigen test (RAT), and had risk factors for progression to severe disease (immunosuppression due to disease or medication (including HIV-positive status with <200 CD4), hemodialysis, cystic fibrosis, age ≥ 65 (molnupiravir period) or age ≥ 75 (nirmatrelvir/ritonavir period), age ≥ 60 or 65 plus a chronic comorbidity, age < 60 or 65 plus 2 chronic comorbidities for molnupiravir or nirmatrelvir/ritonavir periods, respectively. Physicians had to apply for eligible patients through an electronic platform which incorporated eligibility criteria, during the first 3 days of symptom onset or test positivity.

In the analysis patients aged 65 years or older were included, with 4,240 receiving molnupiravir and 13,861 nirmatrelvir/ritonavir.

Regarding the circulation of the different Omicron subvariants, BA.1* was the dominant subgroup until early March 2022, when BA.2* became the most prevalent subgroup that remained dominant until the end of May, when it had been gradually replaced by BA.5* subgroup [24]. The BA.2* circulated at higher proportions during the period of molnupiravir use and BA.5* dominated after the end of May 2022, suggesting that in the period of nirmatrelvir/ritonavir subscription both subvariants were present, but the virus surge observed during June and July was due to the BA.5* subvariants (http://www.eody.gov.gr).

### Study design

Our analysis was performed using a matched retrospective cohort study for the estimation of effectiveness of molnupiravir and nirmatrelvir/ritonavir in preventing hospital admission or death. Outpatient oral antiviral users and controls were matched for age and calendar week of SARS-CoV-2 diagnosis (within the same ISO week). The matching was performed separately for molnupiravir and nirmatrelvir/ritonavir recipients.

On the day 28 after prescription with molnupiravir or nirmatrelvir/ritonavir, medication adherence and treatment side effects were assessed by asking prescribing physicians to fill a questionnaire in the same application platform about the number of pills remaining at the end of the 5-day treatment and adverse effects reported by patients. The questionnaire included the following options about the number of pills missed: i) none, ii) ≤ 5, iii) ≤ 12, iv) > 12, v) all. Patients were classified into two categories, poor adherence including those with all or ≥ 12 pills missed, and complete adherence including those with none pills missed. Adverse drug reactions were reported during the 28 days period of follow-up after the prescription of antivirals. Prescribing physicians were reminded by SMS to fill the questionnaire but their contribution was on a voluntary basis

Demographic data, time of infection, previous SARS-CoV-2 infection, and vaccination status for the study population were available from the covid-19 national registry and the SARS-CoV-2 surveillance data of the National Public Health Organization (NPHO) in Greece.

### Outcomes

The main outcomes of interest were: i) hospital admission with COVID-19 within a period of 10 days following a positive SARS-CoV-2 test, with no admission to ICU, or intubation (symptomatic disease), ii) death from covid-19 within 35 days following a positive SARS-CoV-2 test, and iii) hospitalization, including ICU admission/intubation, or death from covid-19 within 35 days following a positive SARS-CoV-2 test.

### Ethical approval

The study was approved by the Data Protection Officer (DPO) of the Ministry of Health and the ethical committee of the NPHO in Greece.

### Statistical analysis

Continuous variables were described using median and interquartile range (IQR), and for categorical variables frequencies and percentages were used. Factors associated with different outcomes (hospital admission, or covid-19 death, or the combined outcome related to hospital admission, including ICU admission/intubation, or death) were estimated by multivariable logistic regression analysis. Multivariable logistic regression analysis was used to identify each drug’s relative risk (odds ratio, OR) for symptomatic disease or death or the combined outcome, for patients on treatment versus the controls adjusted for age, previous SARS-CoV-2 infection, vaccination status, and time from vaccination. The relative effectiveness of the antivirals was estimated for nirmatrelvir/ritonavir versus molnupiravir recipients adjusted for age, previous SARS-CoV-2 infection, vaccination status, recency of vaccination and co-morbidities (i.e. obesity, BMI >=35; cardiovascular disease, diabetes mellitus, chronic liver disease, chronic kidney disease, and chronic lung disease). Specifically, the number of comorbidities was included in the model as a four-level independent variable (i.e., one, two, three, or more than three) after the exclusion of individuals with moderate or severe immunodeficiency. The effect of treatment adherence in treatment effectiveness was investigated for molnupiravir and nirmatrelvir/ritonavir recipients by comparing patients with complete versus poor or incomplete adherence, adjusted for age, previous SARS-CoV-2 infection, vaccination status, and recency of vaccination.

The drug’s effectiveness was estimated using the reverse of the OR for symptomatic disease or death or the combined outcome for those on molnupiravir or nirmatrelvir/ritonavir versus the controls. Odds ratios (ORs) and 95% confidence intervals (95% CIs) are reported. The level of significance was set at 0.05. All the analyses were conducted using R software.

## Results

Our study population for the estimation of the molnupiravir effectiveness consisted of 4,240 eligible molnupiravir recipients undertaken in outpatient services and 4,240 non-recipients (controls), matched for age and calendar period for SARS-CoV-2 infection. The characteristics of the study population are shown in Table 1. Most of the patients had an age ≥ 80 years (40.87%) and were vaccinated (86.0%). Specifically, 78.4% and 0.8% had received one and two booster doses, respectively, and 6.8% had completed the basic vaccination scheme. Most of the study population was recently vaccinated (≤ 6 months) at the time of SARS-CoV-2 infection (Table 1). The vaccination status and time from most recent dose were almost identical among the controls (Table 1). The proportions of the outcomes for the molnupiravir recipients and non-recipients are also shown in Table 1. Among the recipients, 3.51% (N=149), 0.42% (N=18), and 1.20% (N=51) presented with symptomatic, severe disease (ICU/intubation without death), and death associated with covid-19, respectively, while the percentages for symptomatic disease (8.42%; N=357) and death (3.40%; N=144) were higher among the non-molnupiravir recipients. The co-morbidities for the cases are shown in Table 2, with the most frequent being cardiovascular disease (46.3%), followed by diabetes mellitus (19.1%) and obesity (BMI ≥35) (10.3%). Regarding the self-reported pills missing, 75.6% reported that none of the pills were missing (complete adherence), 8.3% and 2.2% reported ≤ 5 pills and ≤ 12 pills missing, respectively (Table 2). Moreover, 4.1% and 9.8% reported >12 pills and all pills missing (poor adherence) (Table 2). Adverse drug reactions were reported for 3.8% among molnupiravir recipients, with gastrointestinal effects being the most frequently reported (Supplementary Table 1).

**Table 1.**
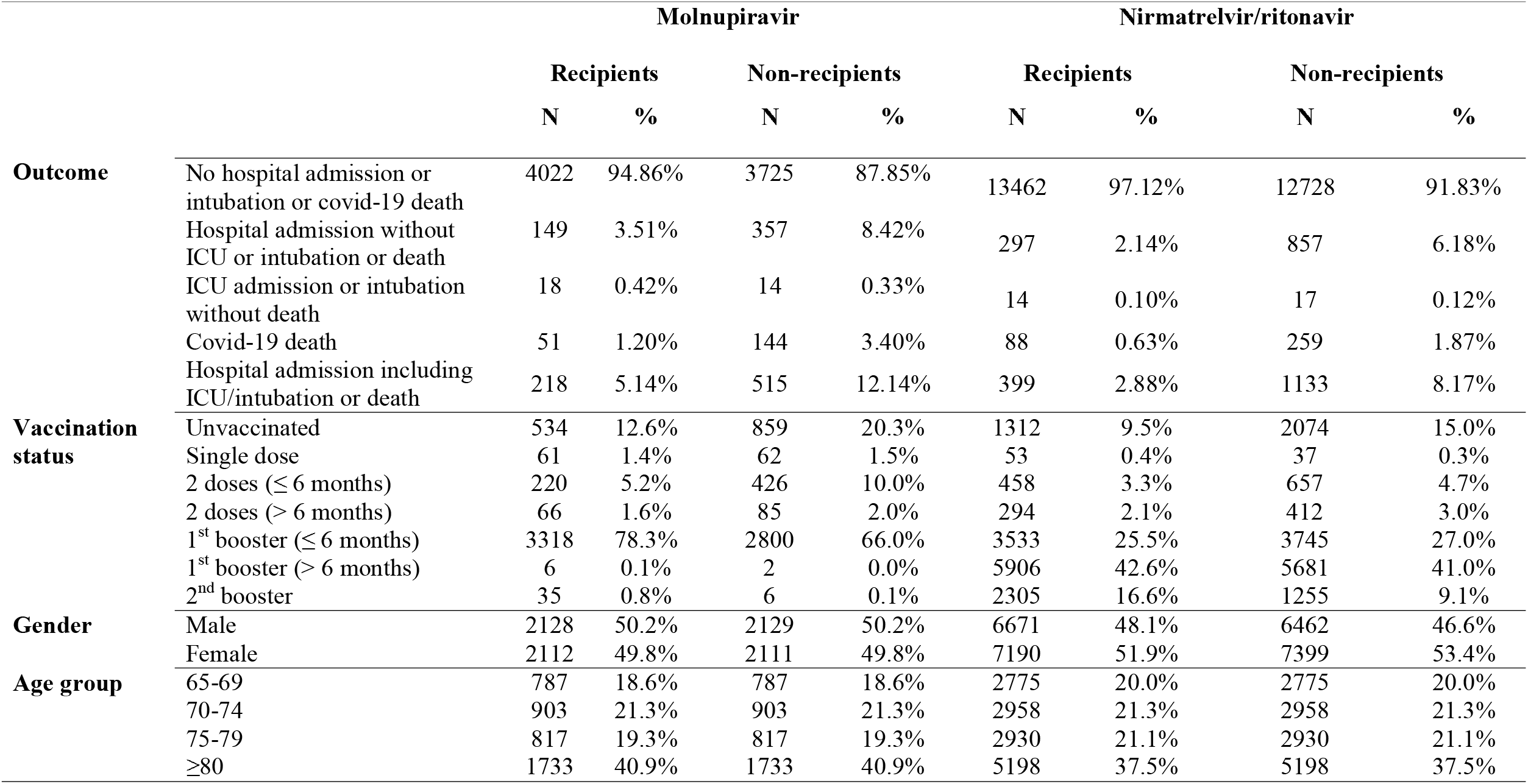
Characteristics of the study population.

**Table 2.** Co-morbidities and treatment adherence for the antiviral’s recipients.

| **Co-morbidities** | **Molnupiravir recipients** |  | **Nirmatrelvir/ritonavir recipients** |  |
| --- | --- | --- | --- | --- |
|  | **N** | **%** | **N** | **%** |
| BMI $\geq 35$ | 435 | 10.26% | 2250 | 16.23% |
| Cardiovascular disease | 1964 | 46.32% | 9668 | 69.75% |
| Moderate or severe immunosuppression | 323 | 7.62% | 1209 | 8.72% |
| DM (diabetes mellitus) | 808 | 19.06% | 3827 | 27.61% |
| Chronic liver disease | 16 | 0.38% | 51 | 0.37% |
| Chronic kidney disease | 239 | 5.64% | 598 | 4.31% |
| Chronic lung disease | 201 | 4.74% | 1046 | 7.55% |
| **Treatment adherence (pills missing)** | **N** | **%** | **N** | **%** |
| None | 3207 | 75.7% | 2626 | 75.01%<sup>1</sup> |
| $\leq 5$ pills | 353 | 8.3% | 308 | 8.80%<sup>1</sup> |
| $\leq 12$ pills | 92 | 2.2% | 141 | 4.03%<sup>1</sup> |
| $> 12$ pills | 172 | 4.1% | 117 | 3.34%<sup>1</sup> |
| All | 416 | 9.8% | 309 | 8.83%<sup>1</sup> |
| Unknown | 0 | 0.0% | 10360 | 74.7% |

<sup>1</sup> Percentages were estimated among individuals with non-missing information about the pills missing

The study population for nirmatrelvir/ritonavir effectiveness analysis included 13,861 eligible nirmatrelvir/ritonavir recipients and 13,861 matched non-recipients. The characteristics of the study population are shown in Table 1. Similarly, to patients treated with molnupiravir, most of the nirmatrelvir/ritonavir recipients had an age ≥ 80 years (37.5%) and were vaccinated with the primary scheme (90.2%) (Table 1). Specifically, 68.1% and 16.6% had received one and two booster doses, respectively; proportions similar to the corresponding ones among the non-recipients (Table 1). Contrary to patients on molnupiravir, those on nirmatrelvir/ritonavir had received the first booster shot earlier, and, therefore, most of the cases receiving a single booster dose had been vaccinated for more than 6 months (42.6%) at the time of infection (Table 1). Among the nirmatrelvir/ritonavir recipients, 2.14% (N=297), 0.1% (N=14), and 0.63% (N=88) developed symptomatic, severe disease, and died from covid-19, respectively, while the proportions for symptomatic disease (6.18%; N=857) and death (1.87%; N=259) were higher among the nirmatrelvir/ritonavir non-recipients (Table 1). The co-morbidities for the recipients are shown in Table 2, with cardiovascular disease (69.8%), diabetes mellitus (27.6%), and obesity (BMI ≥35) (16.2%) being the most frequent and at higher percentages than the non-recipients (Table 2). Regarding the self-reported adherence, data for nirmatrelvir/ritonavir were available for 25.6% of the population (Table 2). Among those with self-reporting information about adherence (N=3,501), 75.0% reported that none of the pills were missing (complete adherence), 8.8% and 4.0% reported ≤ 5 pills and ≤ 12 pills missing, respectively, and 3.3% and 8.8% reported >12 pills and all pills missing (poor adherence) (Table 2). Among those with available data (N=3,274: Supplementary Table 1) on adverse drug reactions, 5.4.% reported adverse drug reactions, with gastrointestinal effects being the most frequently reported (1.03%). (Supplementary Table 1).

The results of multivariable logistic regression analyses for molnupiravir are presented in detail in Table 3. Statistical analysis revealed that the relative risk for symptomatic disease (model 1: OR = 0.40, p < 0.001) or death (models 2: OR = 0.31, p < 0.001) was significantly lower for the recipients (Table 3). The relative risk for hospital admission was lower for recently vaccinated individuals (≤ 6 months) (OR = 0.36, p < 0.001) and for those with previous SARS-CoV-2 infection (OR = 0.45, p = 0.05), whereas it increased with age (OR = 1.06, per year; p < 0.001). No effect was detected for the study population of non-recently vaccinated probably due to the small number of individuals in this group. Similar results were found for death (model 2, Table 3), with recently vaccinated people having lower (OR = 0.36, p < 0.001) and older individuals having higher (OR = 1.11, per year; p < 0.001) relative risk for death. No effect was detected for previous infection and non-recent vaccination on this model (model 2, Table 3). The relative risk for the combined outcome of hospitalization including ICU admission/intubation, or death was similar to hospital admission (OR = 0.40, p<0.001, Supplementary Table 2; model 1, Table 3) with recent vaccination, age and previous infection to have also an effect. To investigate if the treatment effect was more pronounced in specific age groups, we performed a separate analysis in different age groups (models 3-6, Table 3) for the outcome of symptomatic disease. The analysis revealed that treatment with molnupiravir was effective in older ages, and specifically in the age group of 75-79, and in individuals aged ≥80 years. The relative risk for the disease decreased with age in the treatment group (models 4b, 5, and 6, Table 3). For the age group of 70-74, when the two groups of vaccinated people were merged, the effect of treatment approached the borderline of significance. In all age-specific models, vaccination was associated with a reduced relative risk for disease, while no effect was found for previous infection, when included in the models. Including adherence in the model for only patients on molnupiravir treatment, revealed that those with complete adherence had a significantly lower risk for hospitalization (OR = 0.27, p<0.001) compared to people with poor or incomplete adherence, (Table 4; model 1). A similar effect was found for death (OR = 0.24, p<0.001; Table 4; model 2), suggesting that the effectiveness of the drug was approximately 70% higher among patients with complete adherence compared to molnupiravir recipients with partial or poor adherence.

**Table 3.**
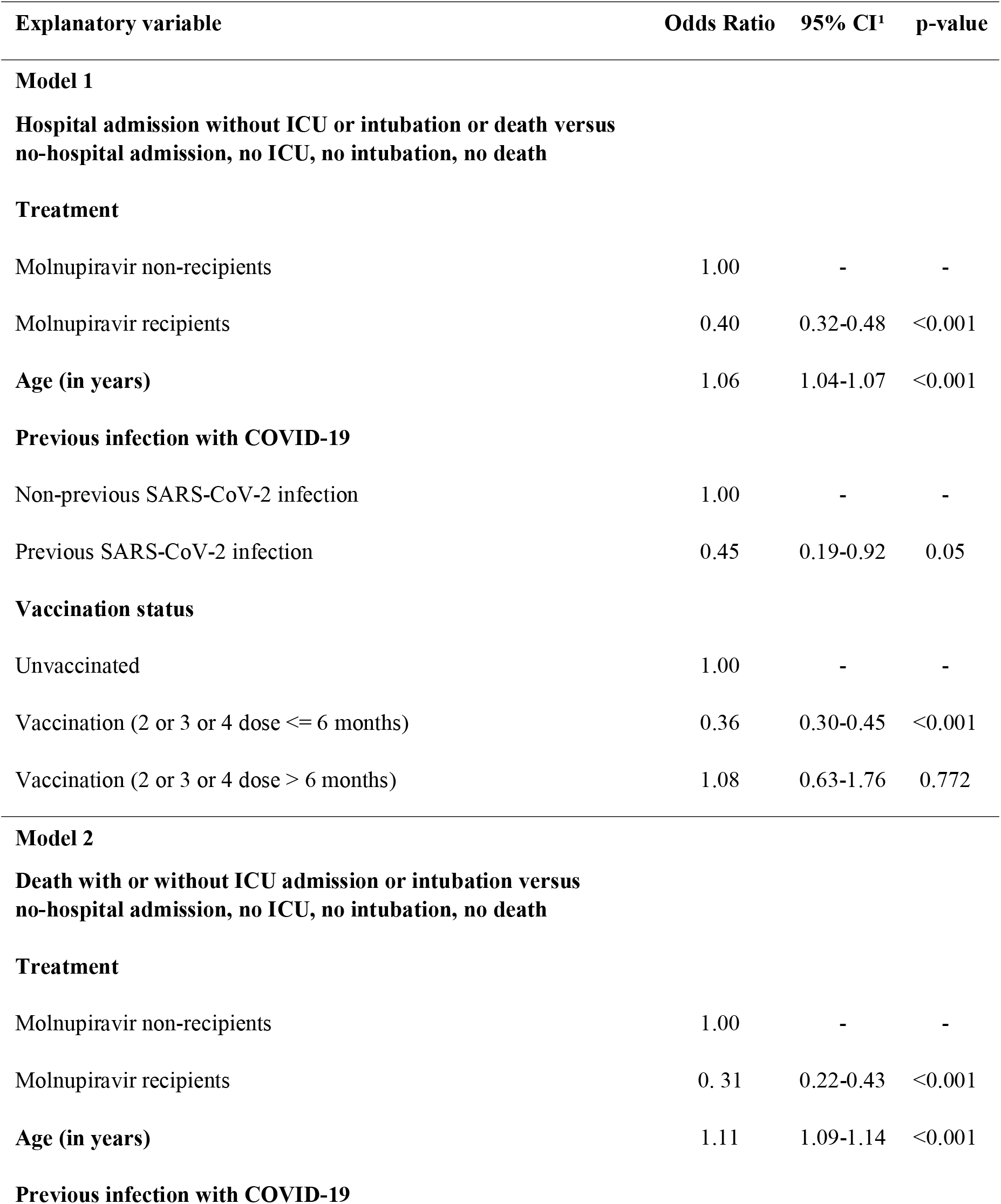
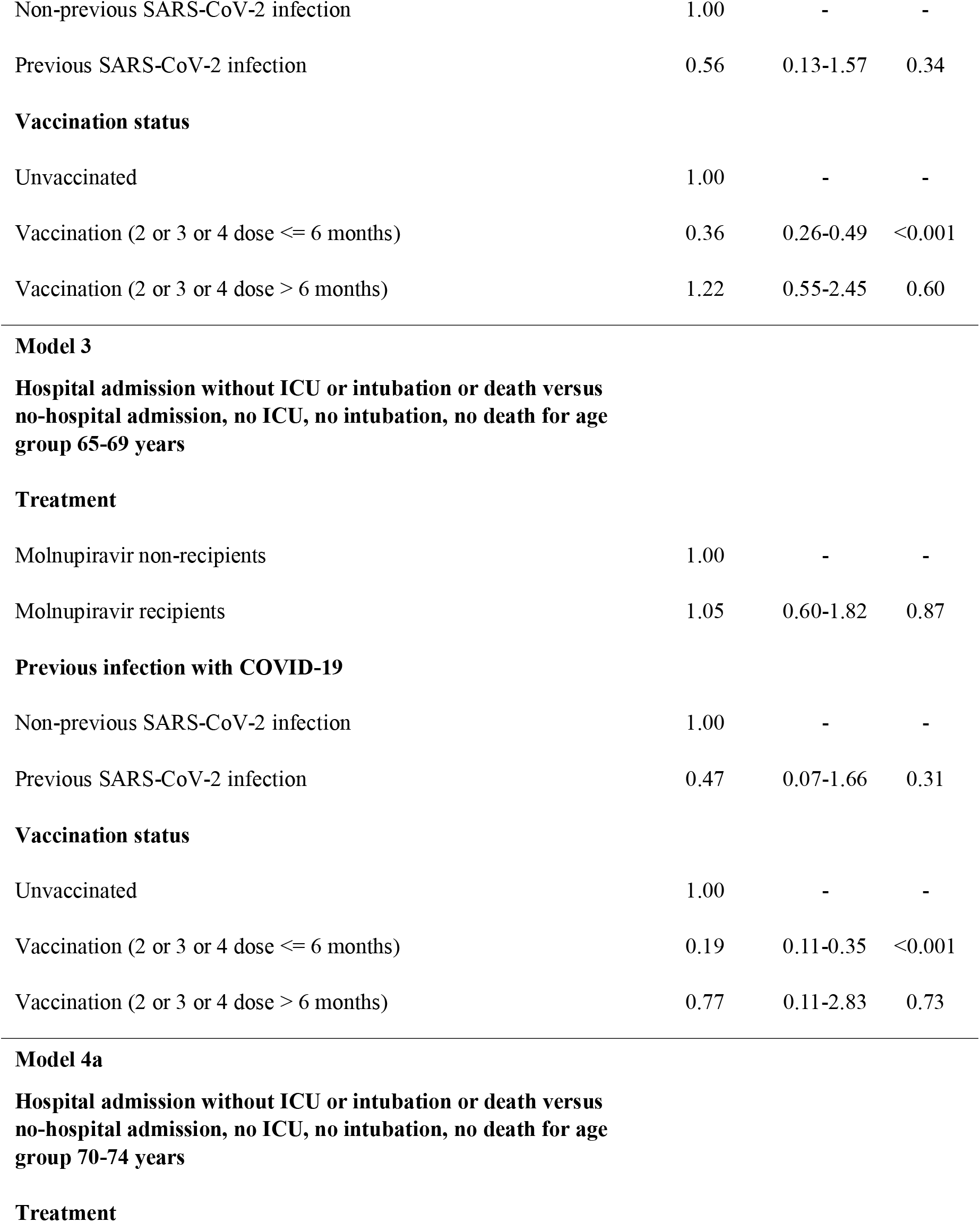
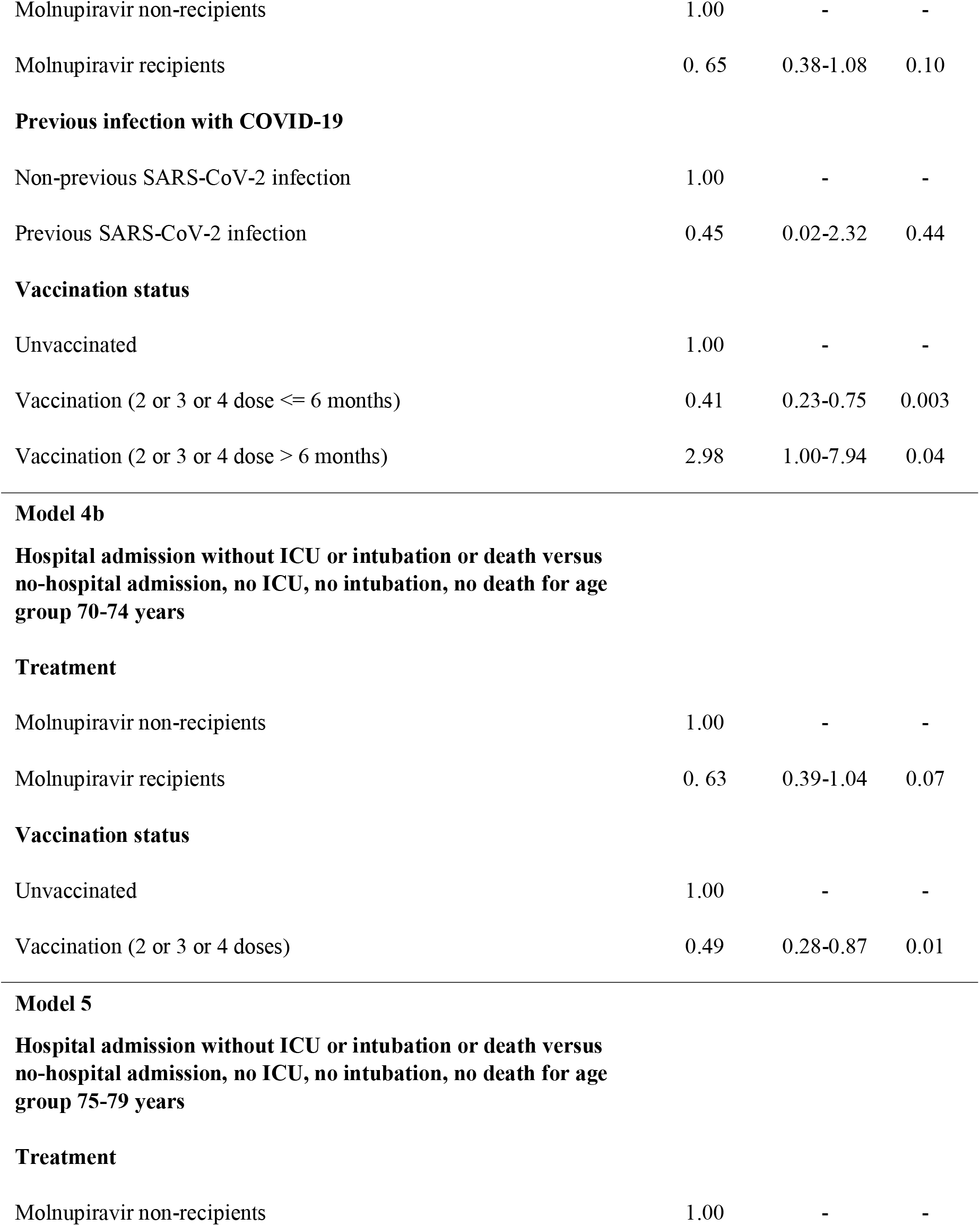
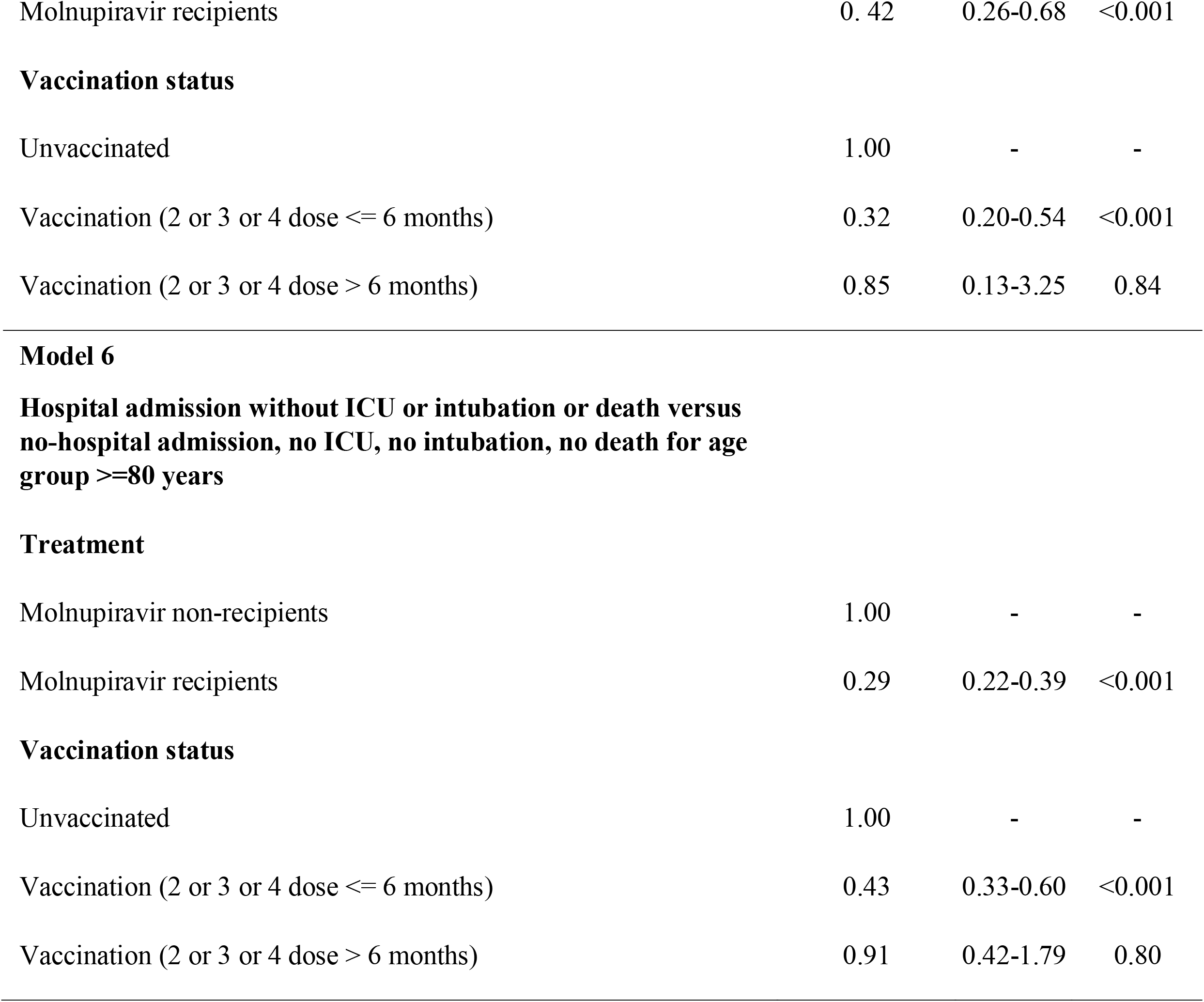
Multivariable logistic regression analysis for the effectiveness of molnupiravir treatment.

| **Explanatory variable** | **Odds Ratio** | **95% CI<sup>1</sup>** | **p-value** |
| --- | --- | --- | --- |
| **Model 1** |  |  |  |
| **Hospital admission without ICU or intubation or death versus no-hospital admission, no ICU, no intubation, no death** |  |  |  |
| **Treatment** |  |  |  |
| Molnupiravir non-recipients | 1.00 | - | - |
| Molnupiravir recipients | 0.40 | 0.32-0.48 | <0.001 |
| **Age (in years)** | 1.06 | 1.04-1.07 | <0.001 |
| **Previous infection with COVID-19** |  |  |  |
| Non-previous SARS-CoV-2 infection | 1.00 | - | - |
| Previous SARS-CoV-2 infection | 0.45 | 0.19-0.92 | 0.05 |
| **Vaccination status** |  |  |  |
| Unvaccinated | 1.00 | - | - |
| Vaccination (2 or 3 or 4 dose ≤ 6 months) | 0.36 | 0.30-0.45 | <0.001 |
| Vaccination (2 or 3 or 4 dose > 6 months) | 1.08 | 0.63-1.76 | 0.772 |
| **Model 2** |  |  |  |
| **Death with or without ICU admission or intubation versus no-hospital admission, no ICU, no intubation, no death** |  |  |  |
| **Treatment** |  |  |  |
| Molnupiravir non-recipients | 1.00 | - | - |
| Molnupiravir recipients | 0.31 | 0.22-0.43 | <0.001 |
| **Age (in years)** | 1.11 | 1.09-1.14 | <0.001 |
| **Previous infection with COVID-19** |  |  |  |
| Non-previous SARS-CoV-2 infection | 1.00 | - | - |
| Previous SARS-CoV-2 infection | 0.56 | 0.13-1.57 | 0.34 |
| **Vaccination status** |  |  |  |
| Unvaccinated | 1.00 | - | - |
| Vaccination (2 or 3 or 4 dose <= 6 months) | 0.36 | 0.26-0.49 | <0.001 |
| Vaccination (2 or 3 or 4 dose > 6 months) | 1.22 | 0.55-2.45 | 0.60 |
| **Model 3** |  |  |  |
| **Hospital admission without ICU or intubation or death versus no-hospital admission, no ICU, no intubation, no death for age group 65-69 years** |  |  |  |
| **Treatment** |  |  |  |
| Molnupiravir non-recipients | 1.00 | - | - |
| Molnupiravir recipients | 1.05 | 0.60-1.82 | 0.87 |
| **Previous infection with COVID-19** |  |  |  |
| Non-previous SARS-CoV-2 infection | 1.00 | - | - |
| Previous SARS-CoV-2 infection | 0.47 | 0.07-1.66 | 0.31 |
| **Vaccination status** |  |  |  |
| Unvaccinated | 1.00 | - | - |
| Vaccination (2 or 3 or 4 dose <= 6 months) | 0.19 | 0.11-0.35 | <0.001 |
| Vaccination (2 or 3 or 4 dose > 6 months) | 0.77 | 0.11-2.83 | 0.73 |
| **Model 4a** |  |  |  |
| **Hospital admission without ICU or intubation or death versus no-hospital admission, no ICU, no intubation, no death for age group 70-74 years** |  |  |  |
| **Treatment** |  |  |  |
| Molnupiravir non-recipients | 1.00 | - | - |
| Molnupiravir recipients | 0.65 | 0.38-1.08 | 0.10 |
| **Previous infection with COVID-19** |  |  |  |
| Non-previous SARS-CoV-2 infection | 1.00 | - | - |
| Previous SARS-CoV-2 infection | 0.45 | 0.02-2.32 | 0.44 |
| **Vaccination status** |  |  |  |
| Unvaccinated | 1.00 | - | - |
| Vaccination (2 or 3 or 4 dose <= 6 months) | 0.41 | 0.23-0.75 | 0.003 |
| Vaccination (2 or 3 or 4 dose > 6 months) | 2.98 | 1.00-7.94 | 0.04 |
| **Model 4b** |  |  |  |
| **Hospital admission without ICU or intubation or death versus no-hospital admission, no ICU, no intubation, no death for age group 70-74 years** |  |  |  |
| **Treatment** |  |  |  |
| Molnupiravir non-recipients | 1.00 | - | - |
| Molnupiravir recipients | 0.63 | 0.39-1.04 | 0.07 |
| **Vaccination status** |  |  |  |
| Unvaccinated | 1.00 | - | - |
| Vaccination (2 or 3 or 4 doses) | 0.49 | 0.28-0.87 | 0.01 |
| **Model 5** |  |  |  |
| **Hospital admission without ICU or intubation or death versus no-hospital admission, no ICU, no intubation, no death for age group 75-79 years** |  |  |  |
| **Treatment** |  |  |  |
| Molnupiravir non-recipients | 1.00 | - | - |
| Molnupiravir recipients | 0.42 | 0.26-0.68 | <0.001 |
| **Vaccination status** |  |  |  |
| Unvaccinated | 1.00 | - | - |
| Vaccination (2 or 3 or 4 dose ≤ 6 months) | 0.32 | 0.20-0.54 | <0.001 |
| Vaccination (2 or 3 or 4 dose > 6 months) | 0.85 | 0.13-3.25 | 0.84 |
| Molnupiravir non-recipients | 1.00 | - | - |
| Molnupiravir recipients | 0.29 | 0.22-0.39 | <0.001 |
| **Vaccination status** |  |  |  |
| Unvaccinated | 1.00 | - | - |
| Vaccination (2 or 3 or 4 dose ≤ 6 months) | 0.43 | 0.33-0.60 | <0.001 |
| Vaccination (2 or 3 or 4 dose > 6 months) | 0.91 | 0.42-1.79 | 0.80 |

**Table 4.**
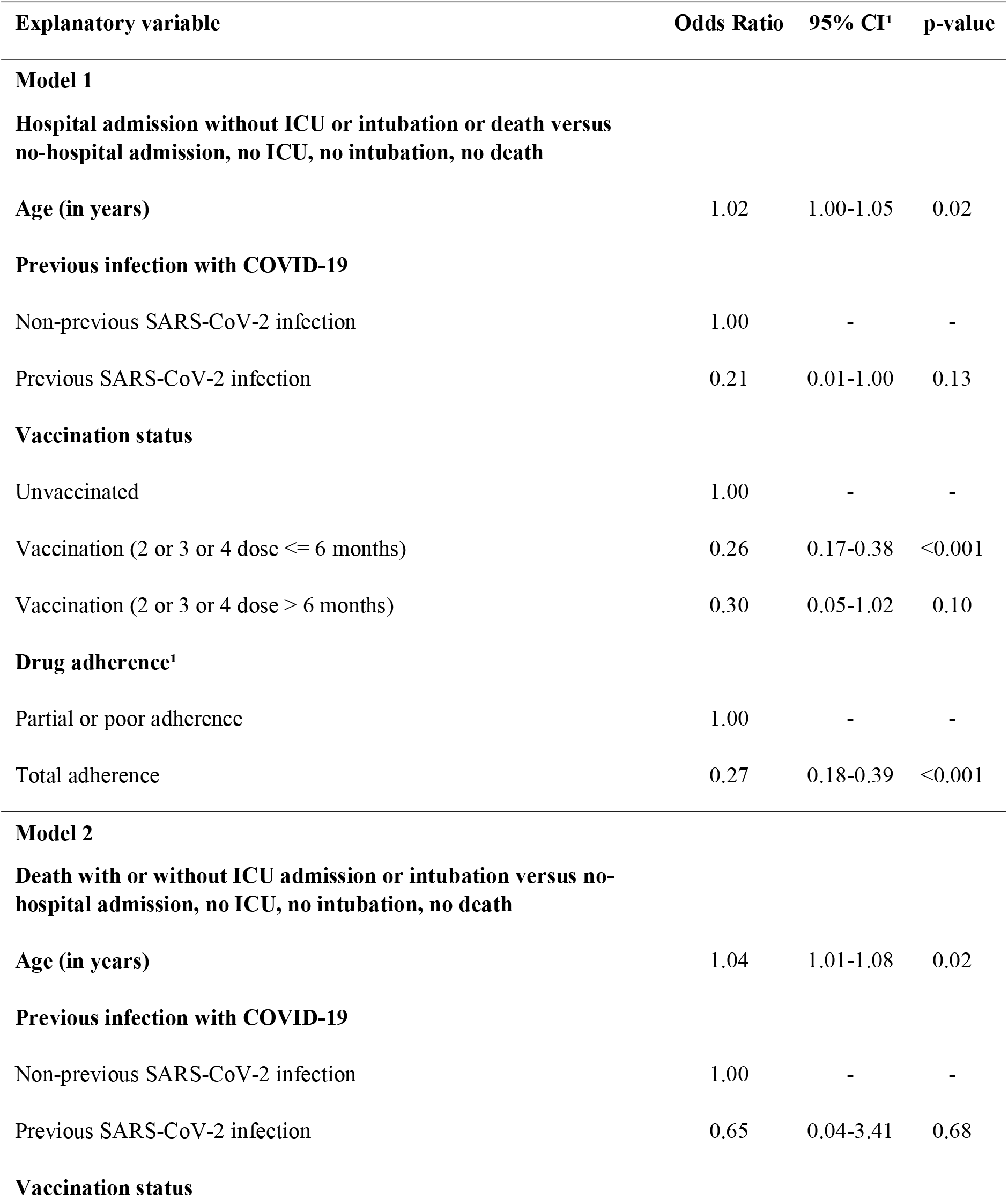
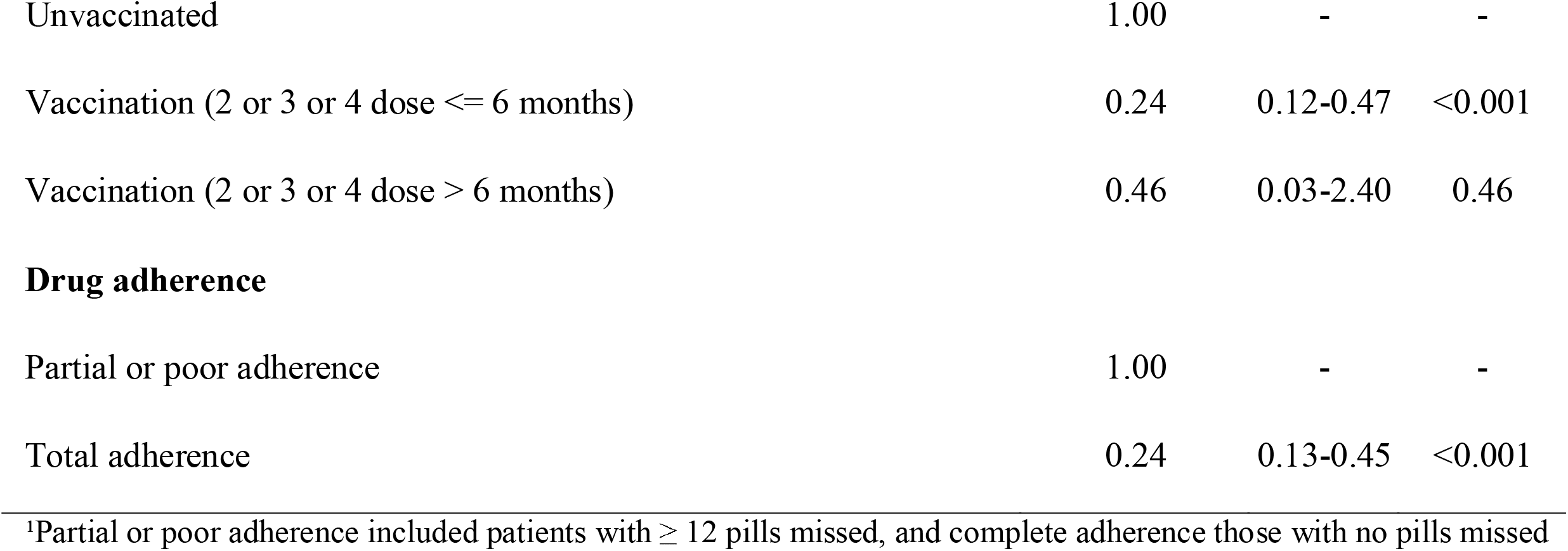
Effectiveness of treatment in the molnupiravir recipients with different levels of adherence.

| **Explanatory variable** | **Odds Ratio** | **95% CI<sup>1</sup>** | **p-value** |
| --- | --- | --- | --- |
| **Model 1** |  |  |  |
| **Hospital admission without ICU or intubation or death versus no-hospital admission, no ICU, no intubation, no death** |  |  |  |
| **Age (in years)** | 1.02 | 1.00-1.05 | 0.02 |
| **Previous infection with COVID-19** |  |  |  |
| Non-previous SARS-CoV-2 infection | 1.00 | - | - |
| Previous SARS-CoV-2 infection | 0.21 | 0.01-1.00 | 0.13 |
| **Vaccination status** |  |  |  |
| Unvaccinated | 1.00 | - | - |
| Vaccination (2 or 3 or 4 dose ≤ 6 months) | 0.26 | 0.17-0.38 | <0.001 |
| Vaccination (2 or 3 or 4 dose > 6 months) | 0.30 | 0.05-1.02 | 0.10 |
| **Drug adherence<sup>1</sup>** |  |  |  |
| Partial or poor adherence | 1.00 | - | - |
| Total adherence | 0.27 | 0.18-0.39 | <0.001 |
| **Model 2** |  |  |  |
| **Death with or without ICU admission or intubation versus no-hospital admission, no ICU, no intubation, no death** |  |  |  |
| **Age (in years)** | 1.04 | 1.01-1.08 | 0.02 |
| **Previous infection with COVID-19** |  |  |  |
| Non-previous SARS-CoV-2 infection | 1.00 | - | - |
| Previous SARS-CoV-2 infection | 0.65 | 0.04-3.41 | 0.68 |
| **Vaccination status** |  |  |  |
| Unvaccinated | 1.00 | - | - |
| Vaccination (2 or 3 or 4 dose ≤ 6 months) | 0.24 | 0.12-0.47 | <0.001 |
| Vaccination (2 or 3 or 4 dose > 6 months) | 0.46 | 0.03-2.40 | 0.46 |
| **Drug adherence** |  |  |  |
| Partial or poor adherence | 1.00 | - | - |
| Total adherence | 0.24 | 0.13-0.45 | <0.001 |

<sup>1</sup>Partial or poor adherence included patients with $\geq 12$ pills missed, and complete adherence those with no pills missed

Overall, the effectiveness of molnupiravir treatment reduced significantly the relative risk for symptomatic disease and death in a real-world evidence study using models adjusted for age, vaccination status, and previous infection. The effect of treatment was more pronounced in older individuals (≥75 years old) with the effectiveness getting higher in individuals aged ≥80 years.

The results of multivariable logistic regression analyses for nirmatrelvir/ritonavir are presented in detail in Table 5. The relative risk for symptomatic disease or death (models 1 and 2, Table 4) was significantly lower for those on treatment (model 1: OR = 0.31, p < 0.001; model 2: OR = 0.28, p < 0.001). The relative risk for hospital admission was lower for individuals with recent (OR = 0.53, p < 0.001) and non-recent (OR = 0.49, p < 0.001) vaccination and for those with previous infection (OR = 0.55, p = 0.003). The relative risk for symptomatic disease, on the other hand, increased with age (OR = 1.09, per year; p < 0.001) (model 1, Table 5). The relative risk for death was significantly lower for patients on treatment (OR = 0.28, p < 0.001), for recently (OR = 0.42, p < 0.001) and non-recently (OR = 0.25, p < 0.001) vaccinated people, and for those with previous infection (OR = 0.32, p = 0.003) (model 2, Table 5), whereas it increased with age (OR = 1.16, per year; p < 0.001) (model 2, Table 5). The relative risk for the combined outcome of hospitalization including ICU admission/intubation, or death was similar to model 1 (OR = 0.32, p<0.001, Supplementary Table 3; model 1, Table 5). In the subsequent analysis for specific age groups, treatment with nirmatrelvir/ritonavir was effective in individuals older than or equal to 70 years (70-74, 75-79, and in people aged ≥80 years) (models 3-6, Table 5). Specifically, the relative risk for symptomatic disease decreased with age in the treatment group (age group 70-74: OR = 0.56, p < 0.001; age group 75-79: OR = 0.39, p < 0.001; age group ≥ 80 years: OR = 0.26, p < 0.001) (models 4-6, Table 5). For all age groups vaccination was associated with a reduced relative risk for hospitalization (models 3-6, Table 5). For nirmatrelvir/ritonavir recipients, with complete adherence had a significantly lower risk for hospitalization (OR = 0.27, p<0.001) compared to people with poor or incomplete adherence, (Table 6; model 1). The findings were similar for death (OR = 0.25, p=0.01; Table 6; model 2).

**Table 5.**
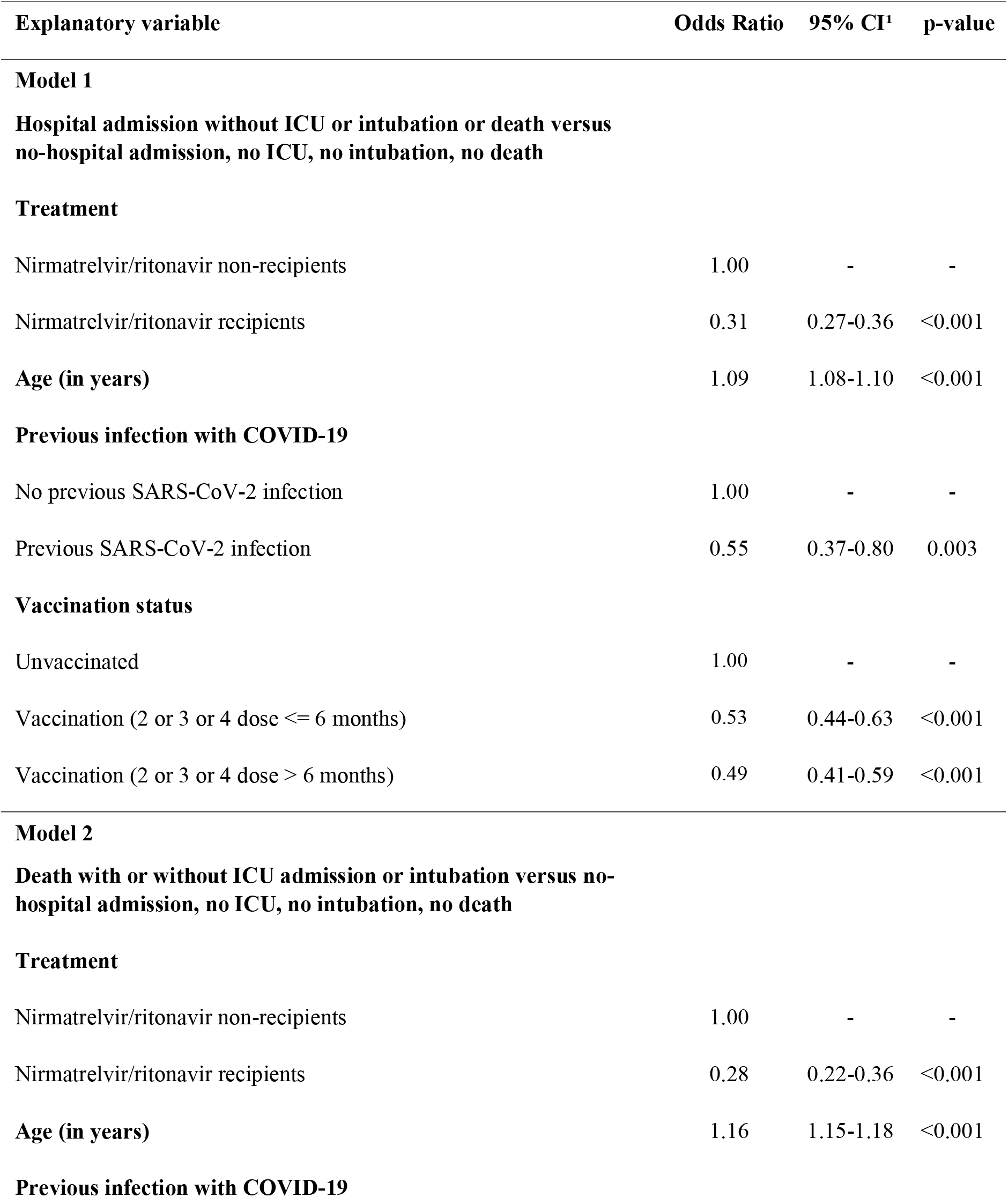
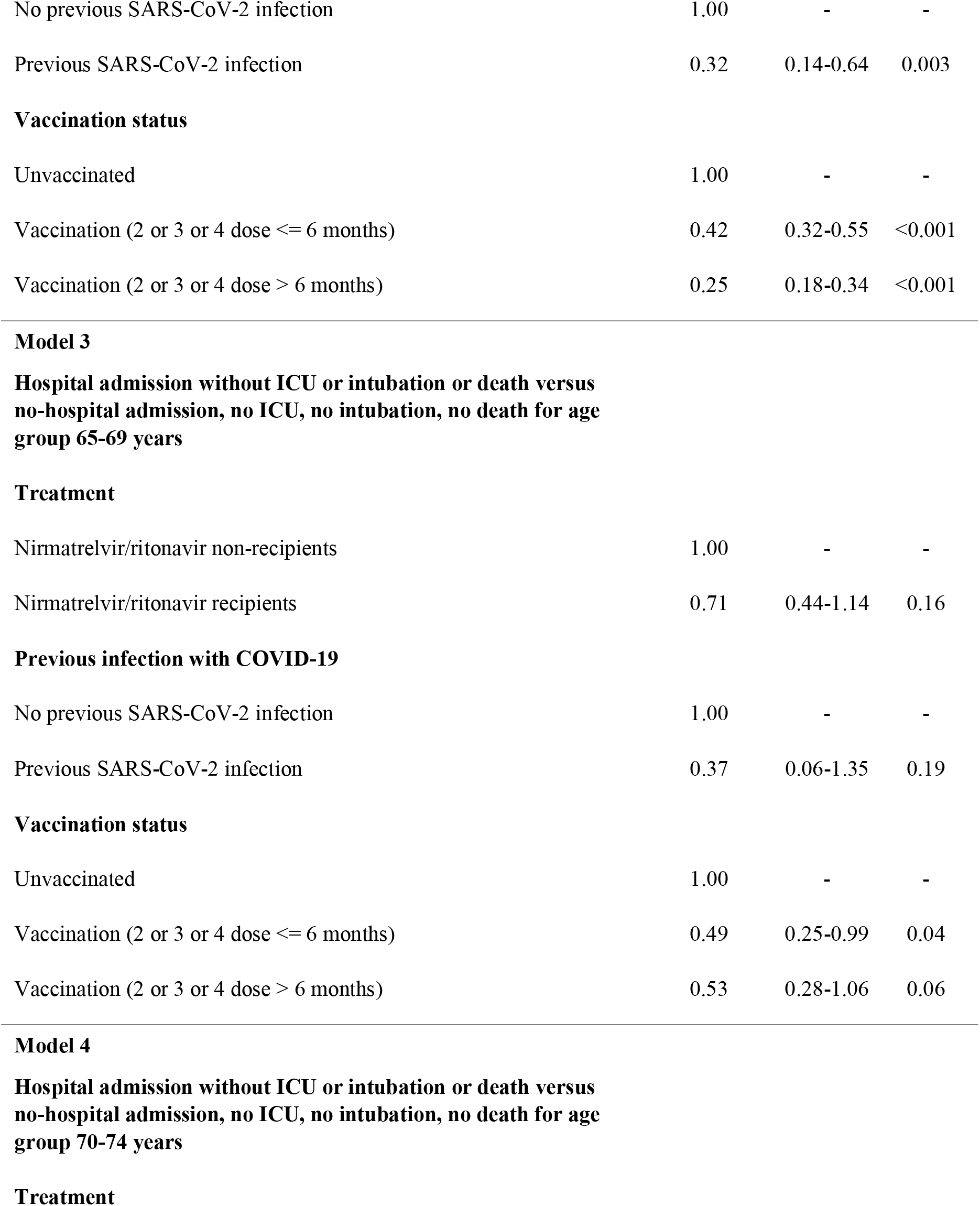
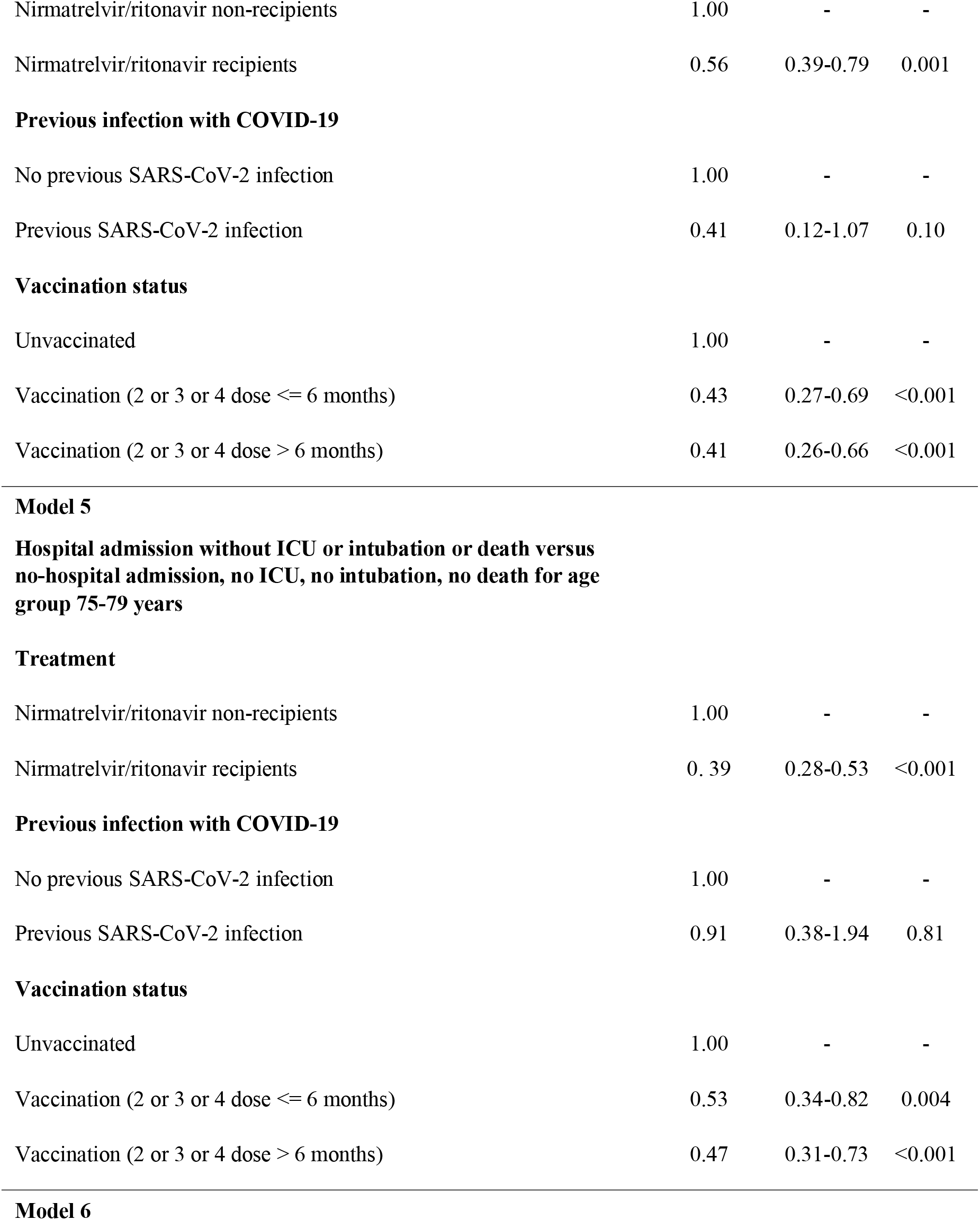
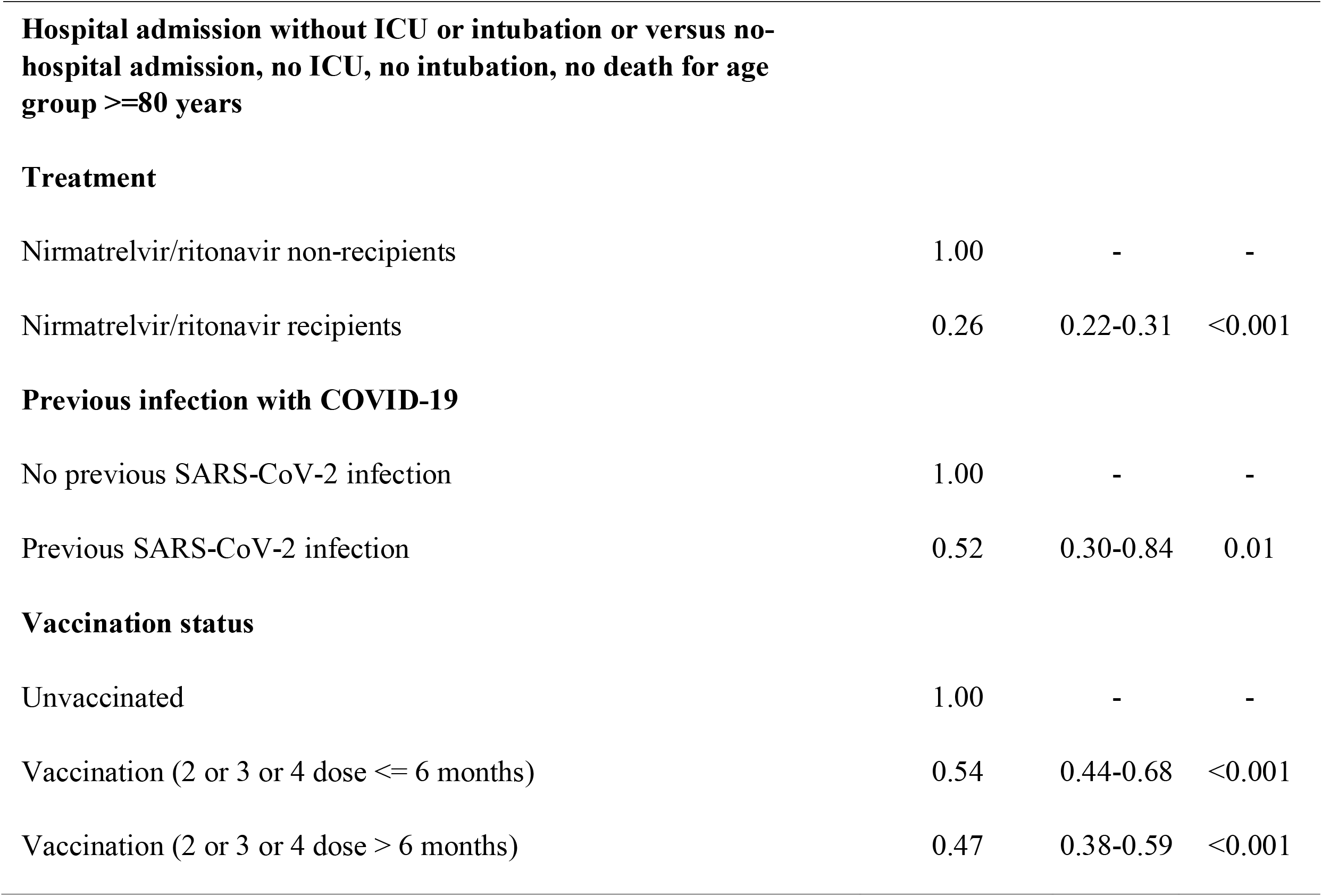
Multivariable logistic regression analysis for the effectiveness of nirmatrelvir/ritonavir treatment.

| **Explanatory variable** | **Odds Ratio** | **95% CI<sup>1</sup>** | **p-value** |
| --- | --- | --- | --- |
| **Model 1** |  |  |  |
| **Hospital admission without ICU or intubation or death versus no-hospital admission, no ICU, no intubation, no death** |  |  |  |
| **Treatment** |  |  |  |
| Nirmatrelvir/ritonavir non-recipients | 1.00 | - | - |
| Nirmatrelvir/ritonavir recipients | 0.31 | 0.27-0.36 | <0.001 |
| **Age (in years)** | 1.09 | 1.08-1.10 | <0.001 |
| **Previous infection with COVID-19** |  |  |  |
| No previous SARS-CoV-2 infection | 1.00 | - | - |
| Previous SARS-CoV-2 infection | 0.55 | 0.37-0.80 | 0.003 |
| **Vaccination status** |  |  |  |
| Unvaccinated | 1.00 | - | - |
| Vaccination (2 or 3 or 4 dose ≤ 6 months) | 0.53 | 0.44-0.63 | <0.001 |
| Vaccination (2 or 3 or 4 dose > 6 months) | 0.49 | 0.41-0.59 | <0.001 |
| **Model 2** |  |  |  |
| **Death with or without ICU admission or intubation versus no-hospital admission, no ICU, no intubation, no death** |  |  |  |
| **Treatment** |  |  |  |
| Nirmatrelvir/ritonavir non-recipients | 1.00 | - | - |
| Nirmatrelvir/ritonavir recipients | 0.28 | 0.22-0.36 | <0.001 |
| **Age (in years)** | 1.16 | 1.15-1.18 | <0.001 |
| **Previous infection with COVID-19** |  |  |  |
| No previous SARS-CoV-2 infection | 1.00 | - | - |
| Previous SARS-CoV-2 infection | 0.32 | 0.14-0.64 | 0.003 |
| **Vaccination status** |  |  |  |
| Unvaccinated | 1.00 | - | - |
| Vaccination (2 or 3 or 4 dose ≤ 6 months) | 0.42 | 0.32-0.55 | <0.001 |
| Vaccination (2 or 3 or 4 dose > 6 months) | 0.25 | 0.18-0.34 | <0.001 |
| **Treatment** |  |  |  |
| Nirmatrelvir/ritonavir non-recipients | 1.00 | - | - |
| Nirmatrelvir/ritonavir recipients | 0.71 | 0.44-1.14 | 0.16 |
| **Previous infection with COVID-19** |  |  |  |
| No previous SARS-CoV-2 infection | 1.00 | - | - |
| Previous SARS-CoV-2 infection | 0.37 | 0.06-1.35 | 0.19 |
| **Vaccination status** |  |  |  |
| Unvaccinated | 1.00 | - | - |
| Vaccination (2 or 3 or 4 dose ≤ 6 months) | 0.49 | 0.25-0.99 | 0.04 |
| Vaccination (2 or 3 or 4 dose > 6 months) | 0.53 | 0.28-1.06 | 0.06 |
| **Treatment** |  |  |  |
| Nirmatrelvir/ritonavir non-recipients | 1.00 | - | - |
| Nirmatrelvir/ritonavir recipients | 0.56 | 0.39-0.79 | 0.001 |
| **Previous infection with COVID-19** |  |  |  |
| No previous SARS-CoV-2 infection | 1.00 | - | - |
| Previous SARS-CoV-2 infection | 0.41 | 0.12-1.07 | 0.10 |
| **Vaccination status** |  |  |  |
| Unvaccinated | 1.00 | - | - |
| Vaccination (2 or 3 or 4 dose ≤ 6 months) | 0.43 | 0.27-0.69 | <0.001 |
| Vaccination (2 or 3 or 4 dose > 6 months) | 0.41 | 0.26-0.66 | <0.001 |
| **Treatment** |  |  |  |
| Nirmatrelvir/ritonavir non-recipients | 1.00 | - | - |
| Nirmatrelvir/ritonavir recipients | 0.39 | 0.28-0.53 | <0.001 |
| **Previous infection with COVID-19** |  |  |  |
| No previous SARS-CoV-2 infection | 1.00 | - | - |
| Previous SARS-CoV-2 infection | 0.91 | 0.38-1.94 | 0.81 |
| **Vaccination status** |  |  |  |
| Unvaccinated | 1.00 | - | - |
| Vaccination (2 or 3 or 4 dose ≤ 6 months) | 0.53 | 0.34-0.82 | 0.004 |
| Vaccination (2 or 3 or 4 dose > 6 months) | 0.47 | 0.31-0.73 | <0.001 |
| Nirmatrelvir/ritonavir non-recipients | 1.00 | - | - |
| Nirmatrelvir/ritonavir recipients | 0.26 | 0.22-0.31 | <0.001 |
| **Previous infection with COVID-19** |  |  |  |
| No previous SARS-CoV-2 infection | 1.00 | - | - |
| Previous SARS-CoV-2 infection | 0.52 | 0.30-0.84 | 0.01 |
| **Vaccination status** |  |  |  |
| Unvaccinated | 1.00 | - | - |
| Vaccination (2 or 3 or 4 dose $\leq 6$ months) | 0.54 | 0.44-0.68 | <0.001 |
| Vaccination (2 or 3 or 4 dose $> 6$ months) | 0.47 | 0.38-0.59 | <0.001 |

**Table 6.**
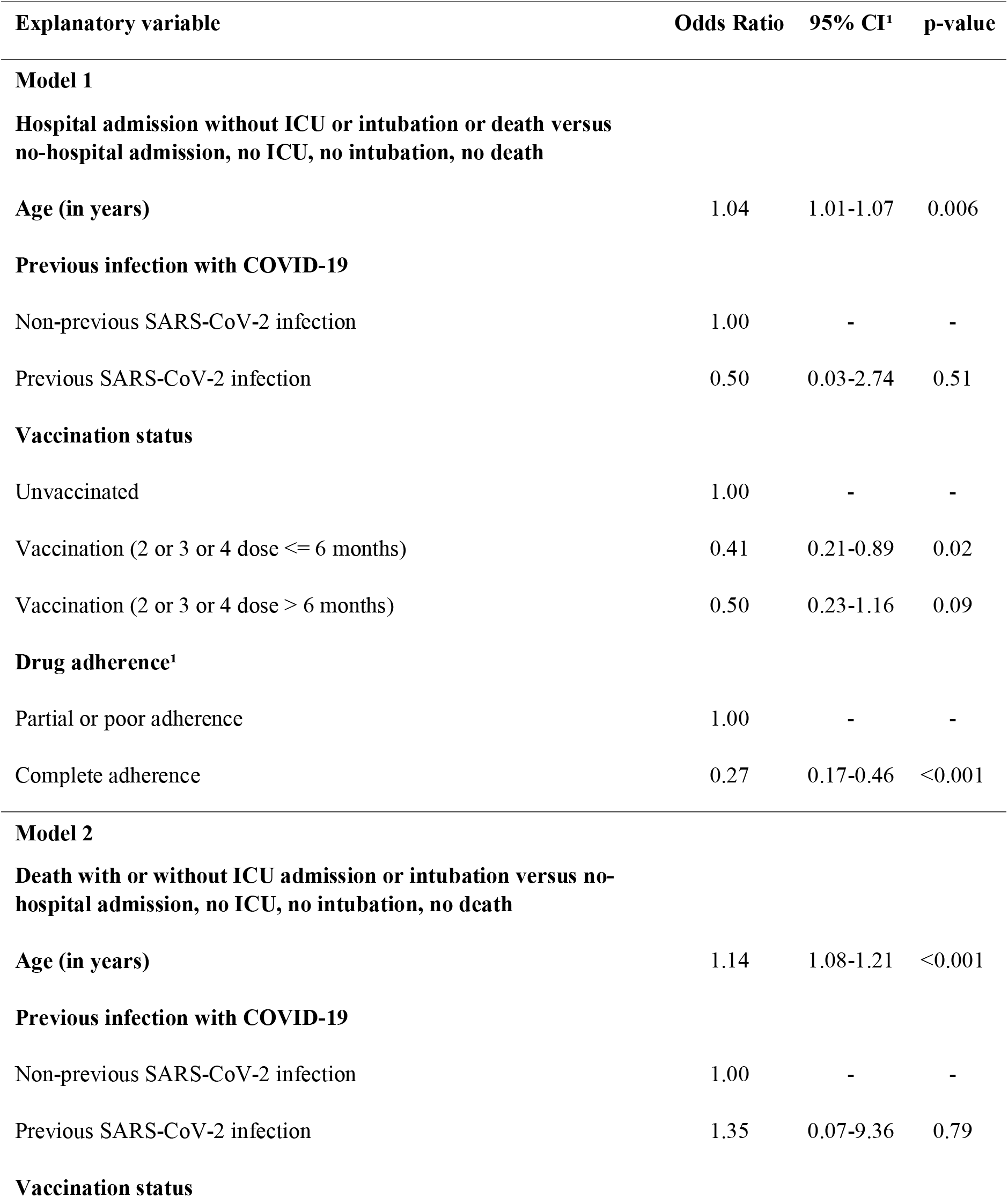
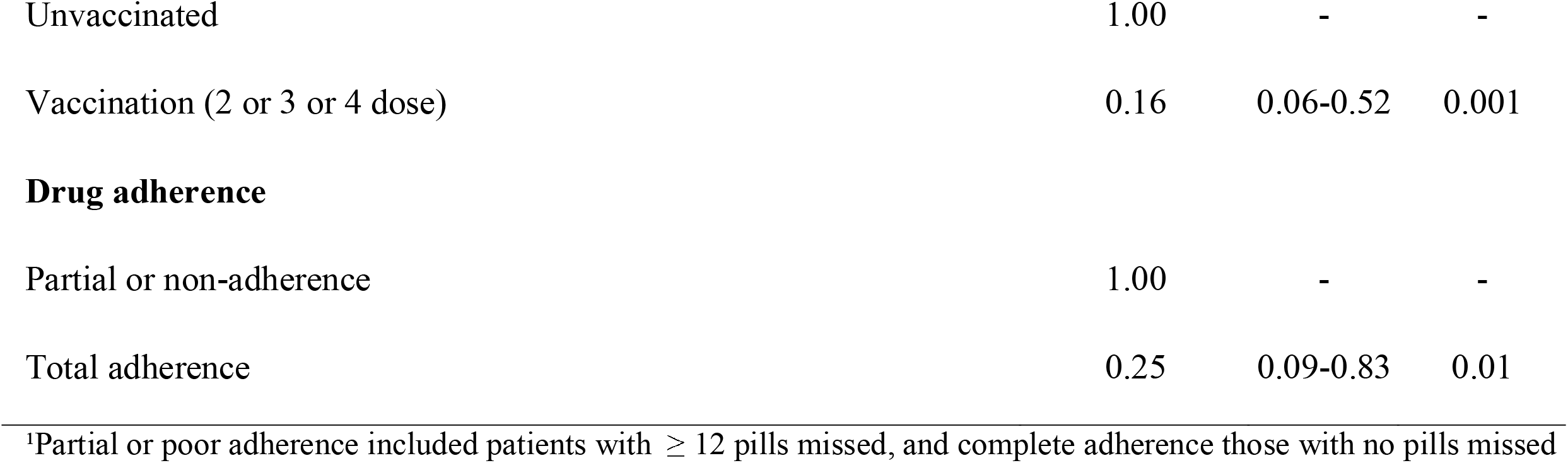
Effectiveness of treatment in nirmatrelvir/ritonavir recipients with different levels of adherence.

| **Explanatory variable** | **Odds Ratio** | **95% CI<sup>1</sup>** | **p-value** |
| --- | --- | --- | --- |
| **Model 1** |  |  |  |
| **Hospital admission without ICU or intubation or death versus no-hospital admission, no ICU, no intubation, no death** |  |  |  |
| **Age (in years)** | 1.04 | 1.01-1.07 | 0.006 |
| **Previous infection with COVID-19** |  |  |  |
| Non-previous SARS-CoV-2 infection | 1.00 | - | - |
| Previous SARS-CoV-2 infection | 0.50 | 0.03-2.74 | 0.51 |
| **Vaccination status** |  |  |  |
| Unvaccinated | 1.00 | - | - |
| Vaccination (2 or 3 or 4 dose ≤ 6 months) | 0.41 | 0.21-0.89 | 0.02 |
| Vaccination (2 or 3 or 4 dose > 6 months) | 0.50 | 0.23-1.16 | 0.09 |
| **Drug adherence<sup>1</sup>** |  |  |  |
| Partial or poor adherence | 1.00 | - | - |
| Complete adherence | 0.27 | 0.17-0.46 | <0.001 |
| **Model 2** |  |  |  |
| **Death with or without ICU admission or intubation versus no-hospital admission, no ICU, no intubation, no death** |  |  |  |
| **Age (in years)** | 1.14 | 1.08-1.21 | <0.001 |
| **Previous infection with COVID-19** |  |  |  |
| Non-previous SARS-CoV-2 infection | 1.00 | - | - |
| Previous SARS-CoV-2 infection | 1.35 | 0.07-9.36 | 0.79 |
| **Vaccination status** |  |  |  |
| Unvaccinated | 1.00 | - | - |
| Vaccination (2 or 3 or 4 dose) | 0.16 | 0.06-0.52 | 0.001 |
| **Drug adherence** |  |  |  |
| Partial or non-adherence | 1.00 | - | - |
| Total adherence | 0.25 | 0.09-0.83 | 0.01 |

<sup>1</sup>Partial or poor adherence included patients with $\geq 12$ pills missed, and complete adherence those with no pills missed

To investigate potential differences between the two groups of treated with different antivirals, we estimated the relative effectiveness of treatment with nirmatrelvir/ritonavir versus molnupiravir. In this analysis, only patients on treatment were included. Statistical analysis revealed that patients who received nirmatrelvir/ritonavir had a lower relative risk for symptomatic disease compared to those on molnupiravir (OR = 0.58, p < 0.001), adjusted for age, previous SARS-CoV-2 infection, vaccination status, and co-morbidities (model 1, Table 7). As in the previous models, vaccination and previous infection reduced the relative risk for hospitalization, while the increasing age and the number of co-morbidities had the opposite effect (model 1, Table 7). The risk for hospital admission increased with the number of comorbidities and specifically patients with two or more comorbidities had a higher risk for hospitalization (model 1, Table 7) compared to those without comorbidities. Comorbidities were associated with a higher risk for hospitalization only in the elderly group of patients (model 6, Table 7). For the age group 65-79 years old, comorbidities were excluded from final analysis since there was no association with the outcome (Table 7). A lower relative risk for death for those on nirmatrelvir/ritonavir was marginally not significant (OR = 0.69, p = 0.09) (model 2, Table 7). A similar effect was found for vaccination, comorbidities, and age, while no significant effect was observed for previous infection. Notably, in the subsequent analyses on specific age groups, the difference in the effectiveness between the two drugs was significant and more pronounced in younger ages (age group 65-69: OR = 0.30, p = 0.002; age group 70-74: OR = 0.39, p = 0.003; ≥80 years old: OR = 0.69, p = 0.03) (models 3, 4, and 6 Table 5). No difference in relative effectiveness was found for people aged between 75-79 years (models 5, Table 5).

**Table 7.**
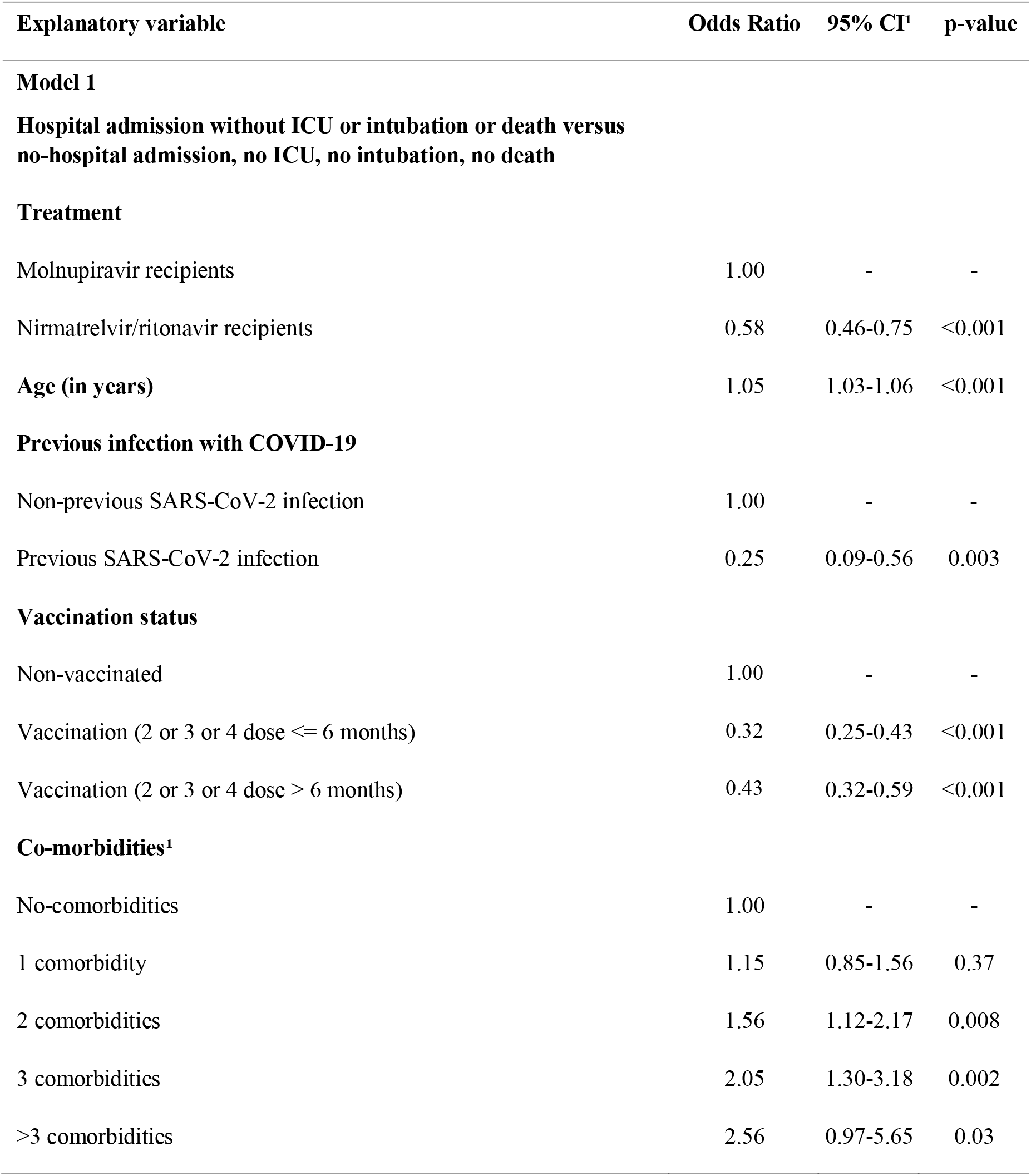
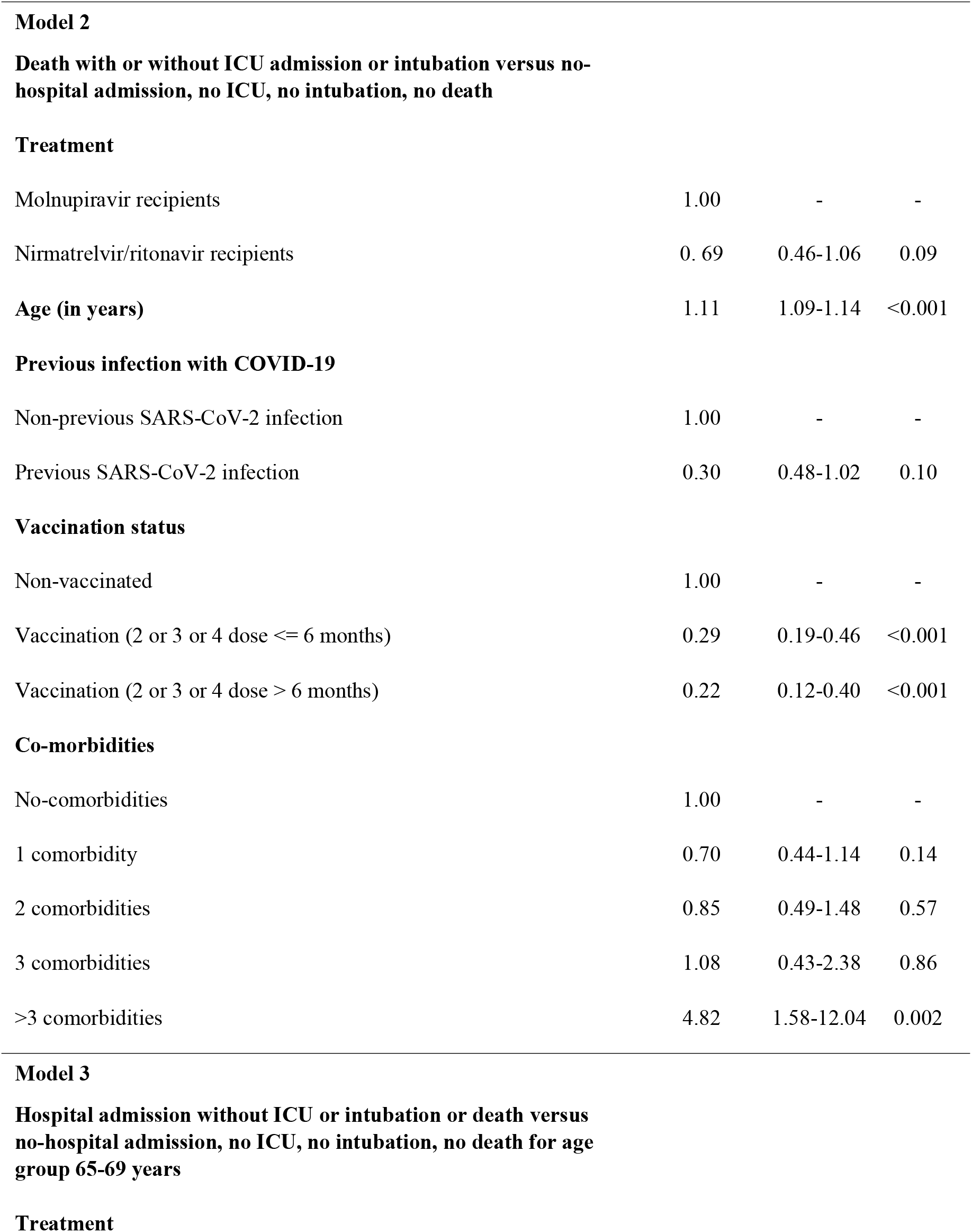
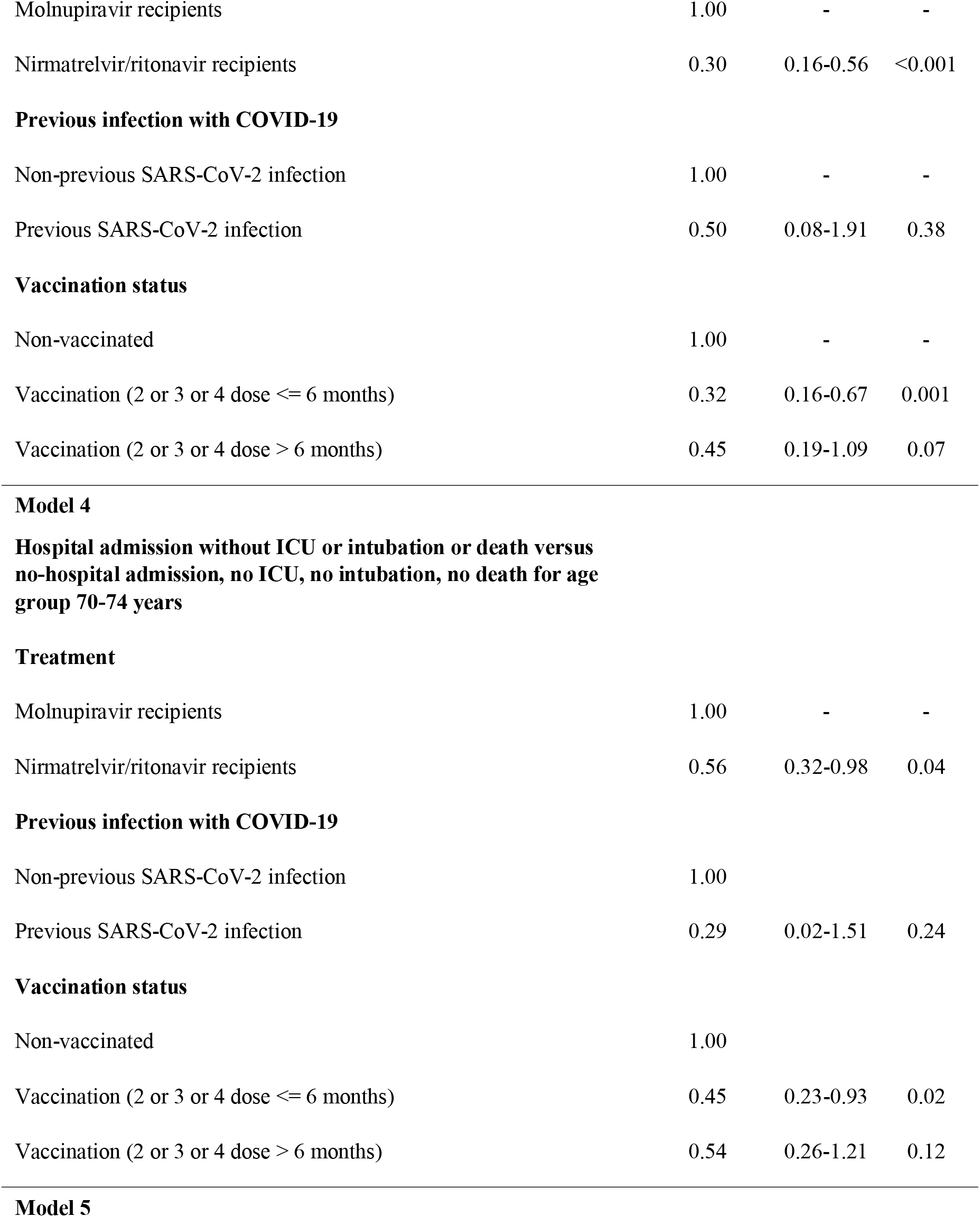
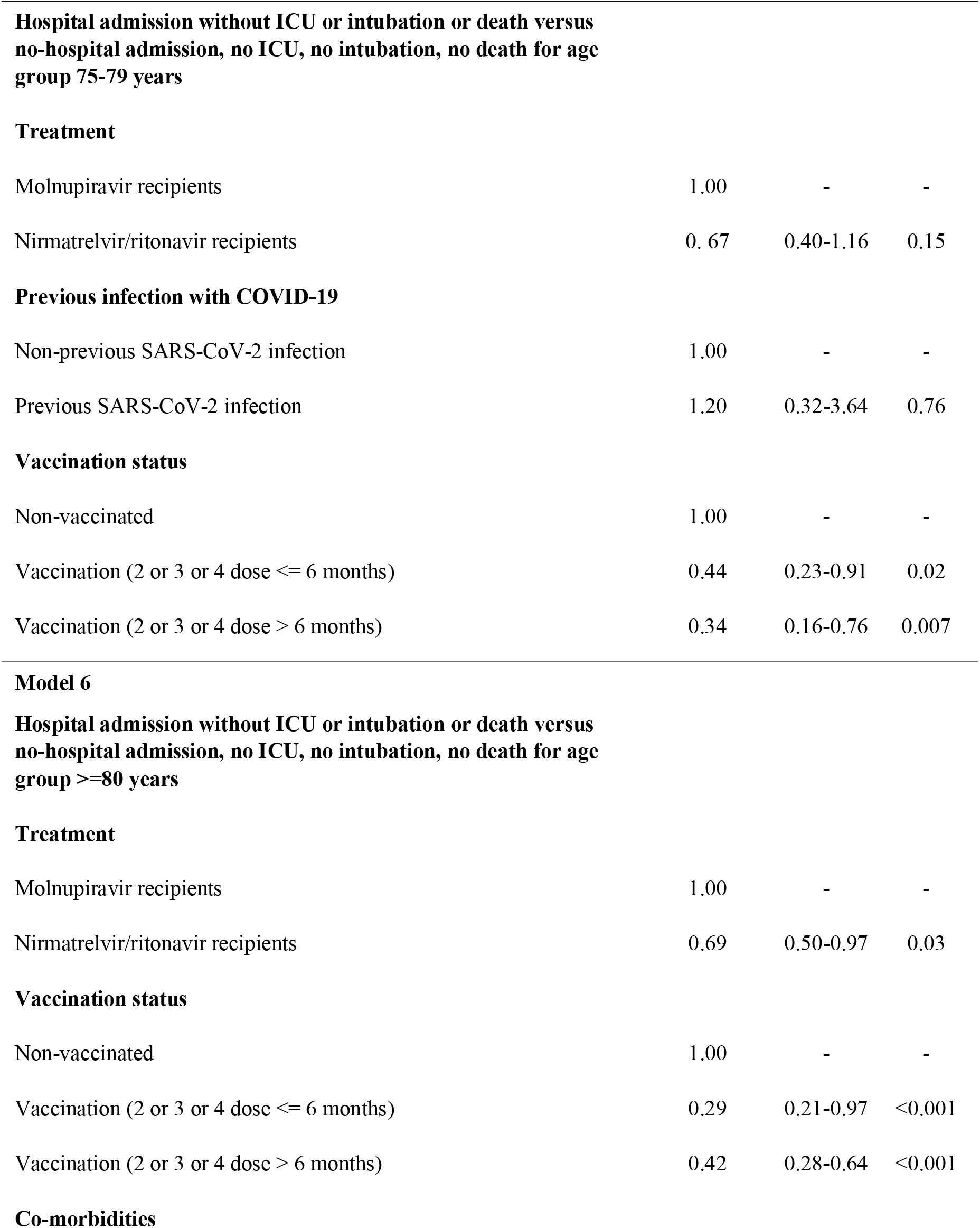
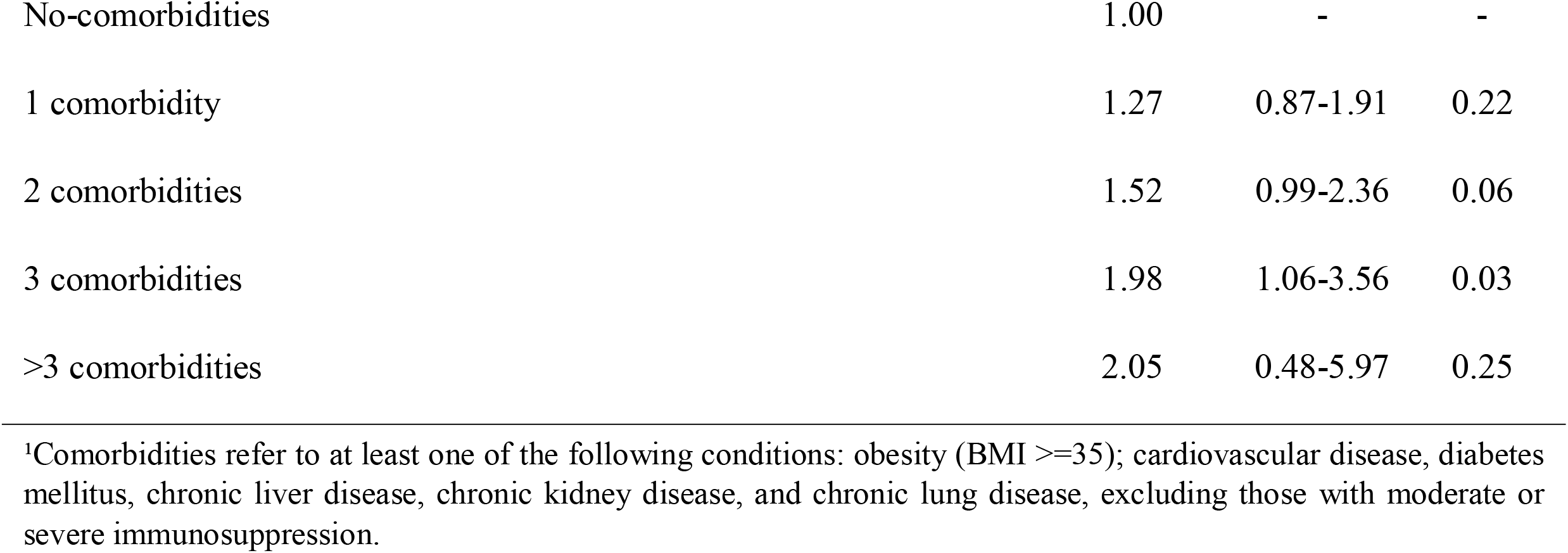
Multivariable logistic regression analysis for the comparison of treatment efficiency between molnupiravir and nirmatrelvir/ritonavir recipients.

| Explanatory variable | Odds Ratio | 95% CI<sup>1</sup> | p-value |
| --- | --- | --- | --- |
| **Model 1** |  |  |  |
| **Hospital admission without ICU or intubation or death versus no-hospital admission, no ICU, no intubation, no death** |  |  |  |
| **Treatment** |  |  |  |
| Molnupiravir recipients | 1.00 | - | - |
| Nirmatrelvir/ritonavir recipients | 0.58 | 0.46-0.75 | <0.001 |
| **Age (in years)** | 1.05 | 1.03-1.06 | <0.001 |
| **Previous infection with COVID-19** |  |  |  |
| Non-previous SARS-CoV-2 infection | 1.00 | - | - |
| Previous SARS-CoV-2 infection | 0.25 | 0.09-0.56 | 0.003 |
| **Vaccination status** |  |  |  |
| Non-vaccinated | 1.00 | - | - |
| Vaccination (2 or 3 or 4 dose ≤ 6 months) | 0.32 | 0.25-0.43 | <0.001 |
| Vaccination (2 or 3 or 4 dose > 6 months) | 0.43 | 0.32-0.59 | <0.001 |
| **Co-morbidities<sup>1</sup>** |  |  |  |
| No-comorbidities | 1.00 | - | - |
| 1 comorbidity | 1.15 | 0.85-1.56 | 0.37 |
| 2 comorbidities | 1.56 | 1.12-2.17 | 0.008 |
| 3 comorbidities | 2.05 | 1.30-3.18 | 0.002 |
| >3 comorbidities | 2.56 | 0.97-5.65 | 0.03 |
| Molnupiravir recipients | 1.00 | - | - |
| Nirmatrelvir/ritonavir recipients | 0.69 | 0.46-1.06 | 0.09 |
| **Age (in years)** | 1.11 | 1.09-1.14 | <0.001 |
| **Vaccination status** |  |  |  |
| Non-vaccinated | 1.00 | - | - |
| Vaccination (2 or 3 or 4 dose ≤ 6 months) | 0.29 | 0.19-0.46 | <0.001 |
| Vaccination (2 or 3 or 4 dose > 6 months) | 0.22 | 0.12-0.40 | <0.001 |
| **Co-morbidities** |  |  |  |
| No-comorbidities | 1.00 | - | - |
| 1 comorbidity | 0.70 | 0.44-1.14 | 0.14 |
| 2 comorbidities | 0.85 | 0.49-1.48 | 0.57 |
| 3 comorbidities | 1.08 | 0.43-2.38 | 0.86 |
| >3 comorbidities | 4.82 | 1.58-12.04 | 0.002 |
| Molnupiravir recipients | 1.00 | - | - |
| Nirmatrelvir/ritonavir recipients | 0.30 | 0.16-0.56 | <0.001 |
| **Previous infection with COVID-19** |  |  |  |
| Non-previous SARS-CoV-2 infection | 1.00 | - | - |
| Previous SARS-CoV-2 infection | 0.50 | 0.08-1.91 | 0.38 |
| **Vaccination status** |  |  |  |
| Non-vaccinated | 1.00 | - | - |
| Vaccination (2 or 3 or 4 dose ≤ 6 months) | 0.32 | 0.16-0.67 | 0.001 |
| Vaccination (2 or 3 or 4 dose > 6 months) | 0.45 | 0.19-1.09 | 0.07 |
| **Treatment** |  |  |  |
| Molnupiravir recipients | 1.00 | - | - |
| Nirmatrelvir/ritonavir recipients | 0.56 | 0.32-0.98 | 0.04 |
| **Vaccination status** |  |  |  |
| Non-vaccinated | 1.00 |  |  |
| Vaccination (2 or 3 or 4 dose ≤ 6 months) | 0.45 | 0.23-0.93 | 0.02 |
| Vaccination (2 or 3 or 4 dose > 6 months) | 0.54 | 0.26-1.21 | 0.12 |
| Molnupiravir recipients | 1.00 | - | - |
| Nirmatrelvir/ritonavir recipients | 0.67 | 0.40-1.16 | 0.15 |
| **Vaccination status** |  |  |  |
| Non-vaccinated | 1.00 | - | - |
| Vaccination (2 or 3 or 4 dose ≤ 6 months) | 0.44 | 0.23-0.91 | 0.02 |
| Vaccination (2 or 3 or 4 dose > 6 months) | 0.34 | 0.16-0.76 | 0.007 |
| Molnupiravir recipients | 1.00 | - | - |
| Nirmatrelvir/ritonavir recipients | 0.69 | 0.50-0.97 | 0.03 |
| **Vaccination status** |  |  |  |
| Non-vaccinated | 1.00 | - | - |
| Vaccination (2 or 3 or 4 dose ≤ 6 months) | 0.29 | 0.21-0.97 | <0.001 |
| Vaccination (2 or 3 or 4 dose > 6 months) | 0.42 | 0.28-0.64 | <0.001 |
| **Co-morbidities** |  |  |  |
| No-comorbidities | 1.00 | - | - |
| 1 comorbidity | 1.27 | 0.87-1.91 | 0.22 |
| 2 comorbidities | 1.52 | 0.99-2.36 | 0.06 |
| 3 comorbidities | 1.98 | 1.06-3.56 | 0.03 |
| >3 comorbidities | 2.05 | 0.48-5.97 | 0.25 |

<sup>1</sup>Comorbidities refer to at least one of the following conditions: obesity (BMI $\geq 35$ ); cardiovascular disease, diabetes mellitus, chronic liver disease, chronic kidney disease, and chronic lung disease, excluding those with moderate or severe immunosuppression.

## Discussion

Our study provides a real-world effectiveness analysis of oral antivirals against SARS-CoV-2 in outpatients at high risk of disease progression. The analysis included all eligible molnupiravir and nirmatrelvir/ritonavir recipients aged ≥ 65 years in Greece, until July 2022, and showed that both drugs reduced the risk of hospitalization and death. Specifically, the risk for hospital admission was significantly lower for molnupiravir recipients, and more specifically in elderly people aged ≥ 75 years. The molnupiravir effectiveness, translated as a reduction in the adjusted OR for hospital admission, was increased with age, with those aged ≥80 years having the highest reduction in the risk for hospitalization across all age groups. Notably, the estimated relative risk for hospitalization was adjusted for vaccination status, and vaccination recency, suggesting that molnupiravir treatment is associated with a reduced risk for disease independently of vaccination. Given that the majority of the study population had received a single booster dose within a time period ≤ 6 months since SARS-CoV-2 infection, our results indicate that molnupiravir treatment reduces the risk for hospitalization even among recent vaccinees with a third dose, and the treatment effect was more pronounced among the elderly population. The adjusted risk for death from COVID-19 was also reduced among molnupiravir recipients versus non-recipients, suggesting that among recently vaccinated people aged ≥65 years, molnupiravir treatment reduced COVID-19 associated mortality. Given that boosted vaccination against SARS-CoV-2 has been shown to provide long-term protection against disease progression and death, our study provides evidence that molnupiravir use further reduced the risk for hospitalization and death. Molnupiravir recipients with complete adherence had a significantly lower risk for hospitalization or death compared to patients with poor or incomplete adherence, suggesting that for high effectiveness of treatment high levels of adherence are necessary.

Nirmatrelvir/ritonavir recipients had similarly a reduced risk of hospital admission or death from COVID-19, versus non-recipients, independently of the vaccination status, recency of vaccination, and previous SARS-CoV-2 infection. For nirmatrelvir/ritonavir the effect of treatment was significant also in the older age groups, specifically those of ≥70 years old with the reduction in the OR for hospitalization being higher among elderly people. For the nirmatrelvir/ritonavir group, non-recent vaccination with a single booster dose was associated with reduced odds of hospitalization or death in contrary to the molnupiravir recipients. Considering that a much higher proportion of patients on nirmatrelvir/ritonavir had been vaccinated 6 months after their infection, this suggests that the non-protective effect of non-recent vaccination in molnupiravir recipients was probably due to the small number of patients in this subgroup. Similarly, to molnupiravir, patients with complete adherence to nirmatrelvir/ritonavir had reduced risk for hospitalization and death compared to those with poor or incomplete levels.

In the comparative analysis between the two drugs, nirmatrelvir/ritonavir recipients had a lower risk of hospitalization and death versus molnupiravir, with the difference being lower for death from COVID-19 versus hospital admission. Notably, nirmatrelvir/ritonavir recipients had a lower risk for hospitalization versus molnupiravir recipients in the youngest age group (65-69) with the difference in the risk being smaller in the age groups of 70-74 and in the group of people aged ≥80 years. The risk for hospitalization or death increased with the number of comorbidities compared to those having reported no health conditions. These findings suggest that the effectiveness of nirmatrelvir/ritonavir was higher than molnupiravir and the difference was more pronounced in the younger ages of our study population. The difference for hospital admission, between the two drugs, remained significant independently of the comorbidities in the drug recipients.

The MOVe-OUT trial has shown that early use of molnupiravir within 5 days of symptoms onset, reduced the relative risk of hospitalization or death by 30% among unvaccinated patients with mild to moderate risk for disease [11]. Similarly, in EPIC-HR trial the relative reduction of hospitalization or death in patients with nirmatrelvir/ritonavir use who were vaccinated with 3 doses at 90.3%, was 88% [13]. Both antivirals have shown a significant reduction in the viral load versus placebo [11, 13], however, the results of MOVe-OUT and EPIC-HR trials have shown that the number needed to treat (NNT) was higher for molnupiravir than nirmatrelvir/ritonavir, suggesting a more robust antiviral effect of the latter [21, 25, 26].

Previous real-world effectiveness studies showed that molnupiravir use was associated with a lower risk for death (HR: 0.76) than non-recipients in Hong Kong, however, the risk for hospitalization was not reduced in drug recipients versus the controls [20]. In the same study, nirmatrelvir/ritonavir recipients had a reduced risk for death (HR:0.34) and hospitalization (HR: 0.76), with mortality risks to be consistently lower among elderly patients with early antiviral use [20]. In a study conducted among U.S. veterans, molnupiravir use reduced the 30-day risk for hospitalization or death among people older than 65 years (RR:0.67) [23]. Similarly, nirmatrelvir-ritonavir recipients had reduced risk for hospitalization or death (RR: 0.53), which was observed mainly in people aged older than 65 years (RR: 0.46) [23]. Notably, no difference was found in the relative risk for hospitalization and death between molnupiravir- and nirmatrelvir/ritonavir-treated patients, but a significant reduction in the absolute risk for death was found for nirmatrelvir/ritonavir recipients versus those with molnupiravir use [23]. In the study by Wai *et al*, both antivirals were associated with reduced risk for hospital admissions [22], and all cause-mortality, with no significant difference to be observed between the two drugs, whereas nirmatrelvir/ritonavir was associated with a stronger effect in hospital admission [22]. In another study by Yip *et al*, only nirmatrelvir/ritonavir was associated with a reduced hazard ratio (WHR: 0.79) for hospitalization, while for molnupiravir no statistically significant difference was observed in the risk for hospitalization versus the non-users [27].

In clinical trials and observational studies, both molnupiravir and nirmatrelvir/ritonavir were associated with reduced risk for death and hospitalization with the effect being more profound among elderly patients [20, 23]. A difference in the effectiveness between the two drugs was not consistently observed [20, 23], with one study detecting a lower absolute risk for death from COVID-19. Our results are consistent with previous findings about the higher reduction in risk for hospitalization and death for both antiviral recipients among elderly people. The reduced risk for hospitalization and death found for nirmatrelvir/ritonavir versus molnupiravir in our setting is also in line with previous findings shown that the reduced risk for hospitalization was more evident for nirmatrelvir/ritonavir users [22].

Our study has some limitations with the most important being the lack of complete data on co-morbidities in the non-recipients of antivirals. To reduce the potential effect of this limitation we selected only drug recipients older than 64 years and age-matched non-recipients (controls). Given that comorbidities such as cardiovascular disease, chronic pulmonary diseases, or diabetes, increase with age, the selection of age-matched controls increase the probability for the controls to have similar co-morbidities with the drug recipients. Another potential limitation is that viral subvariants were untyped in the antiviral’s recipients, however based on the genomic surveillance results, BA.2* dominated during molnupiravir use, and BA.5* took over since the beginning of June suggesting that the majority of cases on nirmatrelvir/ritonavir were infected with BA.5* subvariants.

In conclusion, we provide evidence that the antiviral use significantly reduced the risk for hospitalization and death from COVID-19 in outpatients at high risk of severe disease during the period of BA.2 and BA.5 circulation in Greece. Although that risk for disease progression and death has been significantly reduced due to vaccination, hybrid immunity and the intrinsic characteristics of Omicron (BA.1.1.529) subvariants, we show that in a real-world study, antiviral use further reduced the risk for hospitalization and death with the effect to be more intense in highly vulnerable populations, suggesting that their use in high risk populations is strongly indicated to reduce the burden of disease and adverse outcomes.

## Data Availability

All data produced in the present work are contained in the manuscript

## Author Approval

All authors have seen and approved the manuscript.

## Competing Interests

None

## Funding Statement

This study did not receive any funding

## Notes

### Competing Interest Statement

The authors have declared no competing interest.

### Author Declarations

The study was approved by the Data Protection Officer (DPO) of the Ministry of Health and the ethical committee of the National Public HEalth Organization (NPHO) in Greece.

## References

1. Esper FP, Adhikari TM, Tu ZJ, et al. Alpha to Omicron: Disease Severity and Clinical Outcomes of Major SARS-CoV-2 Variants. J Infect Dis 2022.

2. Fiolet T, Kherabi Y, MacDonald CJ, Ghosn J, Peiffer-Smadja N. Comparing COVID-19 vaccines for their characteristics, efficacy and effectiveness against SARS-CoV-2 and variants of concern: a narrative review. Clin Microbiol Infect 2022; 28:202–21.

3. Asghar N, Mumtaz H, Syed AA, et al. Safety, efficacy, and immunogenicity of COVID-19 vaccines; a systematic review. Immunol Med 2022; 45:225–37.

4. Sharma E, Revinipati S, Bhandari S, et al. Efficacy and Safety of COVID-19 Vaccines-An Update. Diseases 2022; 10.

5. Grana C, Ghosn L, Evrenoglou T, et al. Efficacy and safety of COVID-19 vaccines. Cochrane Database Syst Rev 2022; 12:CD015477.

6. Lee B, Lewis G, Agyei-Manu E, et al. Risk of serious COVID-19 outcomes among adults and children with moderate-to-severe asthma: a systematic review and meta-analysis. Eur Respir Rev 2022; 31.

7. Bobrovitz N, Ware H, Ma X, et al. Protective effectiveness of previous SARS-CoV-2 infection and hybrid immunity against the omicron variant and severe disease: a systematic review and meta-regression. Lancet Infect Dis 2023.

8. Li Z, Liu S, Li F, et al. Efficacy, immunogenicity and safety of COVID-19 vaccines in older adults: a systematic review and meta-analysis. Front Immunol 2022; 13:965971.

9. Beigel JH, Tomashek KM, Dodd LE, et al. Remdesivir for the Treatment of Covid-19 - Final Report. N Engl J Med 2020; 383:1813–26.

10. Gottlieb RL, Vaca CE, Paredes R, et al. Early Remdesivir to Prevent Progression to Severe Covid-19 in Outpatients. N Engl J Med 2022; 386:305–15.

11. Jayk Bernal A, Gomes da Silva MM, Musungaie DB, et al. Molnupiravir for Oral Treatment of Covid-19 in Nonhospitalized Patients. N Engl J Med 2022; 386:509–20.

12. (FDA) UFaDA. Fact sheet for healthcare providers: emergency use authorization for molnupiravir, 2021.

13. Hammond J, Leister-Tebbe H, Gardner A, et al. Oral Nirmatrelvir for High-Risk, Nonhospitalized Adults with Covid-19. N Engl J Med 2022; 386:1397–408.

14. Agency UMaHpR. Oral COVID-19 antiviral, Paxlovid, approved by UK regulator [media release], 2021.

15. Prfizer. Pfizer receives U.S. FDA Emergency Use Authorization for novel COVID-19 oral antiviral treatment [media release], 2021.

16. Agency EM. COVID-19: EMA recommends conditional marketing authorisation for Paxlovid [media release], 2022.

17. Arbel R, Wolff Sagy Y, Hoshen M, et al. Nirmatrelvir Use and Severe Covid-19 Outcomes during the Omicron Surge. N Engl J Med 2022; 387:790–8.

18. Najjar-Debbiny R, Gronich N, Weber G, et al. Effectiveness of Paxlovid in Reducing Severe COVID-19 and Mortality in High Risk Patients. Clin Infect Dis 2022.

19. Vena A, Traman L, Bavastro M, et al. Early Clinical Experience with Molnupiravir for Mild to Moderate Breakthrough COVID-19 among Fully Vaccinated Patients at Risk for Disease Progression. Vaccines (Basel) 2022; 10.

20. Wong CKH, Au ICH, Lau KTK, Lau EHY, Cowling BJ, Leung GM. Real-world effectiveness of molnupiravir and nirmatrelvir plus ritonavir against mortality, hospitalisation, and in-hospital outcomes among community-dwelling, ambulatory patients with confirmed SARS-CoV-2 infection during the omicron wave in Hong Kong: an observational study. Lancet 2022; 400:1213–22.

21. Nyberg T, Ferguson NM, Nash SG, et al. Comparative analysis of the risks of hospitalisation and death associated with SARS-CoV-2 omicron (B.1.1.529) and delta (B.1.617.2) variants in England: a cohort study. Lancet 2022; 399:1303–12.

22. Wai AK, Chan CY, Cheung AW, et al. Association of Molnupiravir and Nirmatrelvir-Ritonavir with preventable mortality, hospital admissions and related avoidable healthcare system cost among high-risk patients with mild to moderate COVID-19. Lancet Reg Health West Pac 2023; 30:100602.

23. Bajema KL, Berry K, Streja E, et al. Effectiveness of COVID-19 treatment with nirmatrelvir-ritonavir or molnupiravir among U.S. Veterans: target trial emulation studies with one-month and six-month outcomes. medRxiv 2022.

24. Kopsidas I, Karagiannidou S, Kostaki EG, et al. Global Distribution, Dispersal Patterns, and Trend of Several Omicron Subvariants of SARS-CoV-2 across the Globe. Trop Med Infect Dis 2022; 7.

25. Dal-Re R, Becker SL, Bottieau E, Holm S. Availability of oral antivirals against SARS-CoV-2 infection and the requirement for an ethical prescribing approach. Lancet Infect Dis 2022; 22:e231–e8.

26. Lee TC, Morris AM, Grover SA, Murthy S, McDonald EG. Outpatient Therapies for COVID-19: How Do We Choose? Open Forum Infect Dis 2022; 9:ofac008.

27. Yip TCF, Lui GCY, Lai MSM, et al. Impact of the use of oral antiviral agents on the risk of hospitalization in community COVID-19 patients. Clin Infect Dis 2022.

