## Supplementary Tables for "Real-world effectiveness of molnupiravir and nirmatrelvir/ritonavir among COVID-19 community, highly vaccinated patients with high risk for severe disease: Evidence that both antivirals reduce the risk for disease progression and death"

**Supplementary Table 1 Adverse drug reactions for molnupiravir and nirmatrelvir/ritonavir recipients**

|  | **Molnupiravir**  **recipients** | | **Νirmatrelvir/ritonavir recipients** | |
| --- | --- | --- | --- | --- |
| **Adverse drug reactions** | **Ν** | **%** | **Ν** | **%** |
| Gastrointestinal (GI) effects | 107 | 2.52% | 143 | 1.03% |
| Allergy | 7 | 0.17% | 3 | 0.02% |
| Headache, dizziness | 22 | 0.52% | 11 | 0.08% |
| Other | 21 | 0.61% | 28 | 0.20% |
| Subtotal | 162 | 3.82% | 185 | 1.33% |
| No adverse drug reactions | 4013 | 94.65% | 3274 | 23.62% |
| Unknown | 65 | 1.53% | 10402 | 75.05% |
| Total | 4240 | 100% | 13861 | 100.0% |

**Supplementary Table 2 Multivariable logistic regression analysis for the effectiveness of molnupiravir treatment**

| **Explanatory variable** | **Odds Ratio** | **95% CI¹** | **p-value** |
| --- | --- | --- | --- |
| **Hospital admission including ICU/intubation or death versus no-hospital admission, no ICU, no intubation, no death** |  |  |  |
| **Treatment** |  |  |  |
| Molnupiravir non-recipients | 1.00 | - | - |
| Molnupiravir recipients | 0.40 | 0.34-0.48 | <0.001 |
| **Age (in years)** | 1.07 | 1.06-1.08 | <0.001 |
| **Previous infection with COVID-19** |  |  |  |
| Non-previous SARS-CoV-2 infection | 1.00 | - | - |
| Previous SARS-CoV-2 infection | 0.45 | 0.19-0.92 | 0.02 |
| **Vaccination status** |  |  |  |
| Unvaccinated | 1.00 | - | - |
| Vaccination (2 or 3 or 4 dose <= 6 months) | 0.34 | 0.29-0.41 | <0.001 |
| Vaccination (2 or 3 or 4 dose > 6 months) | 1.17 | 0.75-1.77 | 0.48 |

**Supplementary Table 3 Multivariable logistic regression analysis for the effectiveness of nirmatrelvir/ritonavir treatment**

| **Explanatory variable** | **Odds Ratio** | **95% CI¹** | **p-value** |
| --- | --- | --- | --- |
| **Hospital admission including ICU/intubation or death versus no-hospital admission, no ICU, no intubation, no death** |  |  |  |
| **Treatment** |  |  |  |
| Molnupiravir non-recipients | 1.00 | - | - |
| Molnupiravir recipients | 0.32 | 0.28-0.36 | <0.001 |
| **Age (in years)** | 1.10 | 1.10-1.11 | <0.001 |
| **Previous infection with COVID-19** |  |  |  |
| Non-previous SARS-CoV-2 infection | 1.00 | - | - |
| Previous SARS-CoV-2 infection | 0.47 | 0.33-0.66 | <0.001 |
| **Vaccination status** |  |  |  |
| Unvaccinated | 1.00 | - | - |
| Vaccination (2 or 3 or 4 dose <= 6 months) | 0.49 | 0.42-0.57 | <0.001 |
| Vaccination (2 or 3 or 4 dose > 6 months) | 0.41 | 0.35-0.48 | <0.001 |
